# Comparative efficacy of Qigong exercise and other external treatments of traditional Chinese medicine on insomnia in the elderly: A network meta-analysis

**DOI:** 10.1101/2025.04.24.25326329

**Authors:** Xinxin Ye, Huanju Liu, Zhuzhu Qin, Yining Tao, Danfeng Chen, Jiaxin Zhang, Xiaofei Liang, Hui Li, Ruizhe Jiang, Ping Wang, Cong Huang

## Abstract

**Introduction:** Insomnia triggers an enormous burden on people’s physical and mental health, especially for the elderly. Traditional Chinese medicine (TCM) therapies were widely used for curing senile insomnia, while the efficiency among various external treatments of TCM (ETs-TCM) was rarely reported.

**Method:** Our network meta-analysis aims to compare and evaluate the optimal ETs-TCM to cure insomnia in the geriatric population. We systematically searched randomized controlled trials (RCTs) published until April 27, 2023 from nine databases. All participants satisfying the diagnosis criteria were recruited without limiting sex, nationality, or race.

**Result:** From the initial 13,846 terms searched, we read 2,936 full-text articles and finally included 85 studies (12,724 participants) in our network meta-analysis. The total effective (TE) rate and Pittsburgh Sleep Quality Index (PSQI) score were selected standards for evaluating the effects of ETs-TCM. Evaluated by TE, Qigong plus acupressure massage has the highest probability (83.5%) of being the most effective treatment from its Surface Under the Cumulative Ranking Curves (SUCRA) at 96.7. Additionally, when evaluated using the PSQI, Acupressure Massage had the highest probability (32.9%) of being the most effective treatment (SUCRA=90.9).

**Conculsion:** Our findings suggest that the combination of acupressure massage and Qigong, or either method alone, may be effective treatments for insomnia in the elderly. This study provides valuable insight for future senile insomnia interventions using ETs-TCM, emphasizing the significance of manual therapy and gentle mind-body exercise as an alternative to medication.

## 1. Introduction

Insomnia is a prevalent sleep disorder that affects a large population, with an estimated incidence rate of 20% among the general public [1]. This disorder is characterized by difficulty falling or staying asleep [2], leading to poor sleep quality and daytime fatigue. As we age, the incidence rate of insomnia increases [3], primarily due to the degeneration of the central nervous system. Studies have shown that nearly 50% of older adults struggle with initiating or maintaining sleep [4], negatively impacting mental and physical health. Commonly observed symptoms in these patients include stress, depression, and cognitive deficits [5]. Furthermore, insomnia has been linked to an increased risk of metabolic and cardiovascular diseases [6, 7], placing a significant burden on individuals and society.

Various therapies have been applied to cure insomnia from different angles, including cognitive-behavioral therapy [8], medications [9], sleep restriction therapy [10], and Traditional Chinese Medicine (TCM) therapies [11], etc. Each of these therapies addresses different aspects of insomnia, such as the underlying causes, symptoms, or maintaining good sleep habits. TCM is a holistic approach to health and processes a unique diagnostic and treatment system used for over 2,000 years [12]. From the angle of TCM, insomnia results from the body’s imbalanced energy flow (*yin* and *yang qi*) [13]. So, their therapies aim to promote balance and restore health to the body and mind, thus improving sleep quality.

TCM uses a range of natural remedies, including herbal medicine, dietary therapy, external treatments, etc. The external treatment of TCM is a specific TCM therapy applied externally to the body or physical exercise. Studies have shown that the external treatment of TCM can improve sleep quality and reduce insomnia symptoms in older adults [14], making it a valuable treatment option for this population. Unlike sleep drugs, external TCM therapies provide a more natural way to cure insomnia without too many side effects or drug resistance [15]. Many are also popular in conjunction with medication to enhance their effectiveness in improving sleep quality, like the acupressure [16] and fire dragon pot [17]. Additionally, gentle exercises such as health Qigong [18], including Tai Chi and others, can offer numerous physical and psychological benefits while improving sleep. Also, some TCM were applied in animal models to test their effectiveness in curing insomnia [19, 20]. However, it is still challenging to evaluate the effectiveness of different external TCM therapies on senile insomnia.

Our study comprehensively evaluated the efficacy of traditional Chinese medicine-based external treatments (ETs-TCM) for insomnia using network meta-analysis. This advanced statistical method synthesizes evidence from multiple clinical trials, enabling simultaneous comparisons of various interventions through both direct and indirect evidence [21]. By estimating the relative efficacy and ranking interventions probabilistically, network meta-analysis provides a powerful framework for guiding evidence-based clinical decisions, especially when direct head-to-head comparisons are unavailable. This study ranks ETs-TCM therapies across multiple standards, offering valuable insights for older adults with insomnia, healthcare professionals, and researchers, while paving the way for future clinical trials to improve treatment outcomes in this population.

## 2. Methods

This systematic review was registered with PROSPERO (CRD 42021278116). Our study followed Preferred Reporting Items for Systematic Reviews and Meta-analyses.

### 2.1. Search Strategy

Our search strategy followed the principle of “PICOS”. The database included: PUBMED, EMbase, Cochrane Library, CINAHL, PsycINFO, Web of Science, CNKI, VIP and Wanfang Data databases. We also tracked related reviews and articles until April 27, 2023. The detailed search strategy of all available databases is presented in Appendix 1.

MeSH terms, keywords, or free words included: “Chronic Insomnia”, “Sleepless”, “Sleeplessness”, “Medicine, Chinese Traditional”, “Complementary Therapies”, “Alternative therapies”, “Auriculotherapy”, “Acupuncture”, “Acupressure”, “Moxibustion”, “Acupoint injection”, “Scraping therapy”, “Cupping therapy”, “Massage”, “Tuina”, “Qigong”, “Tai Ji”, “Ch’i Kung”, “Wuqinxi”, “Baduanjin”, “Liuzijue”, “Breathing exercises”, “Five-element music”, “Acupuncture point paste”, “Chinese herbal soaking” and “Randomized controlled trial”.

### 2.2. Selection Criteria

Included studies met the following 3 criteria: 1) RCTs whether blind or not; 2) patients diagnosed with insomnia in the elderly (≥65 years), regardless of nationality, race, age, and gender; 3) intervention limited to ETs-TCM.

We note that ETs-TCM in criteria 3 included Acupuncture, Combination therapy, Qigong (specifically health Qigong and Tai Chi), Acupuncture and moxibustion, Auricular therapy, Health education, Herbal medicine foot bath, Acupressure massage, Moxibustion, Acupuncture plus acupressure massage, Qigong plus acupressure massage, Manipulative therapy, Chinese herbal fumigation, Aromatherapy, Scrapie, Moxibustion plus acupressure massage, Five-element music, Acupoint sticking therapy, Chinese medicine iontophoresis treatment, patients who underwent TCM with routine treatment (RT). We specifically included "health Qigong" and "Tai Chi" as the types of Qigong exercises being studied, as these are commonly used for health promotion and symptom relief in elderly populations. The following were excluded: animal research articles, repeated or incomplete articles, reviews, case studies, guides, experiences, comments, and registration information. Meanwhile, we excluded RT groups mixed with other designated ETs-TCM patterns.

### 2.3. Data Extraction and Quality Assessment

Data were extracted with Excel and cross-checked by eight evaluators (Huanju Liu, Zhuzhu Qin, Yining Tao, Danfeng Chen, Jiaxin Zhang, Xiaofei Liang, Ruizhe Jiang, and Ping Wang) independently. Evaluators contacted the authors for confirmation when necessary. Disagreements were resolved by sending them to the third evaluator (Xinxin Ye and Cong Huang) to obtain a consensus.

The form included study characteristics (first author’s name, publication year, study setting and design), patient characteristics (sample size, mean age, and sex ratio), interventions (types, duration, and frequency), and outcomes. The risk bias for each article was estimated following the Cochrane Handbook for Systematic Reviews of Interventions (Version 5.1.0) [22].

### 2.4. Outcome Measurement

The outcomes of our network meta-analysis included the total effective (TE) rate and Pittsburgh Sleep Quality Index (PSQI) scores [23]. The TE rate = clinical control rate + markedly effective rate + effective rate [24]. The PSQI is a self-rated questionnaire that measures sleep quality in clinical populations. It consists of seven component scores and the sum (global score). The resulting total scores ranged from 0 to 21. A patient was considered to be experiencing sleep disturbances when the global score ≥ 5.

### 2.5. Statistical Analysis

To explore the effects of various external treatments of TCMs individually from the control group, we performed a pairwise meta-analysis across all outcomes, reporting effect sizes with mean differences (MDs) and 95% confidence intervals (CIs) using post-intervention outcomes by the random-effects model. Whenever possible, we evaluated missing continuous outcome data using the latest follow-up data and treated missing dichotomous outcome data with intention-to-treat. Missing SDs were calculated from p values, t values, CIs, or standard errors [25]. We then conducted the network meta-analysis using Stata software (Version 14.0; StataCorp, College Station, TX, USA) within a frequentist framework [26].

We produced network graphs using the network package and visually presented the results with league tables. We assessed global and local coherence between direct and indirect sources of evidence by comparing the fit and parsimony of consistency and inconsistency models and calculating the difference between direct and indirect estimates in all closed loops in the network [27]. We estimated the ranking probabilities of each intervention at each possible rank and summarized the treatment hierarchy as the surface under the cumulative ranking curve (SUCRA) [28]. Funnel plots were utilized to evaluate the potential presence of publication bias.

## 3. Results

### 3.1. Literature search and study characteristics

We identified a total of 13,846 citations from our initial literature search. After screening the titles and abstracts, 2,936 full-text articles were assessed for eligibility. A total of 2,856 full-text articles were excluded for the following reasons: studies with unavailable data (n = 6), studies that were not randomized controlled trials (RCTs) (n = 75), studies that did not include relevant interventions (n = 475), studies with participants who were not elderly (n = 2,053), studies lacking extractable outcomes (n = 82), duplicate publications (n = 13), and articles for which the full text was not available (n = 158). Ultimately, 85 studies (12,724 participants) were included in our meta-analysis. These studies were published between 2001 and 2023, with 46 trials focusing on efficacy and 60 trials focusing on PSQI scores (as shown in Figure 1). Table 1 presents the characteristics of the included studies. An assessment of bias using the Cochrane Risk of Bias Tool revealed an acceptably low degree of bias across the different parameters scored (see Figure 2 and Appendix 2 for details).

**Figure 1:**
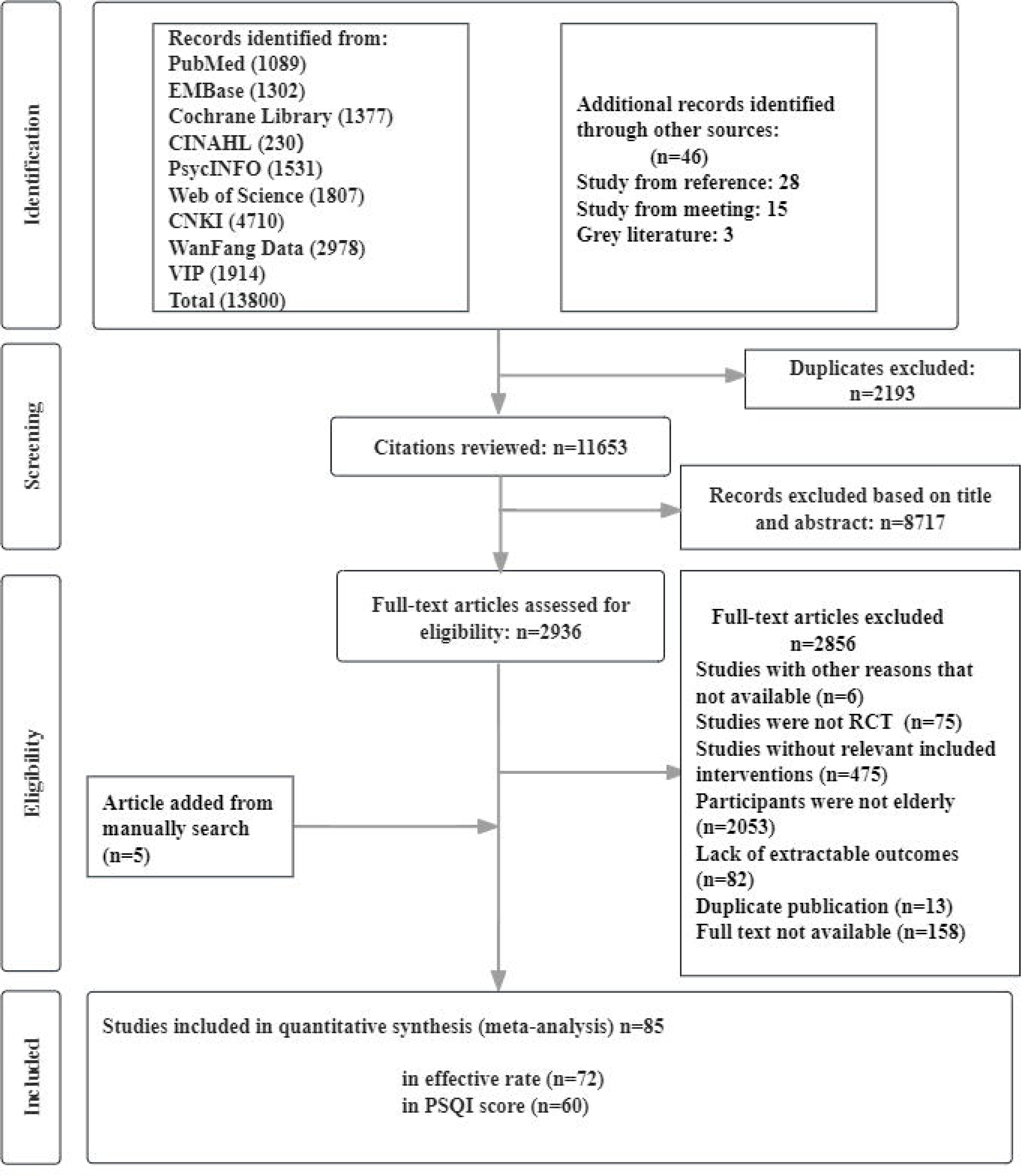
Flowchart illustrating the selection for included studies. Abbreviations: CNKI, China National Knowledge Infrastructure; VIP, Chinese Scientific Journal Database.

**Figure 2:**
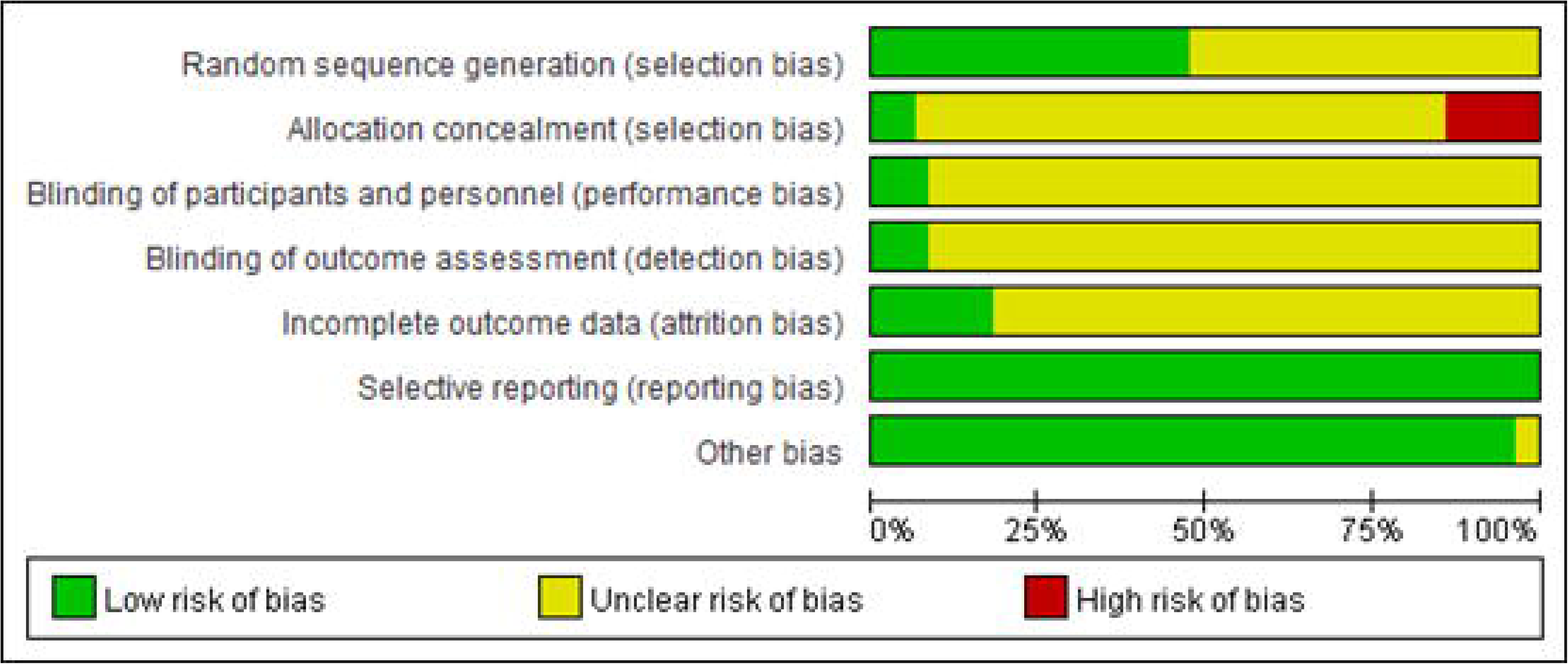
The overall risk of bias for all included studies

### 3.2. TE rate of ETs-TCM for insomnia in the elderly

For the analysis of the TE rate, we included 72 studies with a total of 11,555 participants. Of these studies, 69 adopted a 2-arm design, while three adopted a 3-arm design to compare various ETs-TCM (Appendix 3). A pairwise analysis indicated that ETs-TCM were effective, with an overall I^2^ value of 65% (Appendix 3.1). We also performed an inconsistency test using network analysis, which revealed no significant local inconsistency; the detailed results for this test can be found in Appendix 4.1. Additionally, the funnel plot showed no evidence of publication bias (Appendix 5.1). To further visualize these results, we also created a network plot for the TE rate (Figure 3A), which displays all the available comparisons from the included trials.

**Figure 3:**
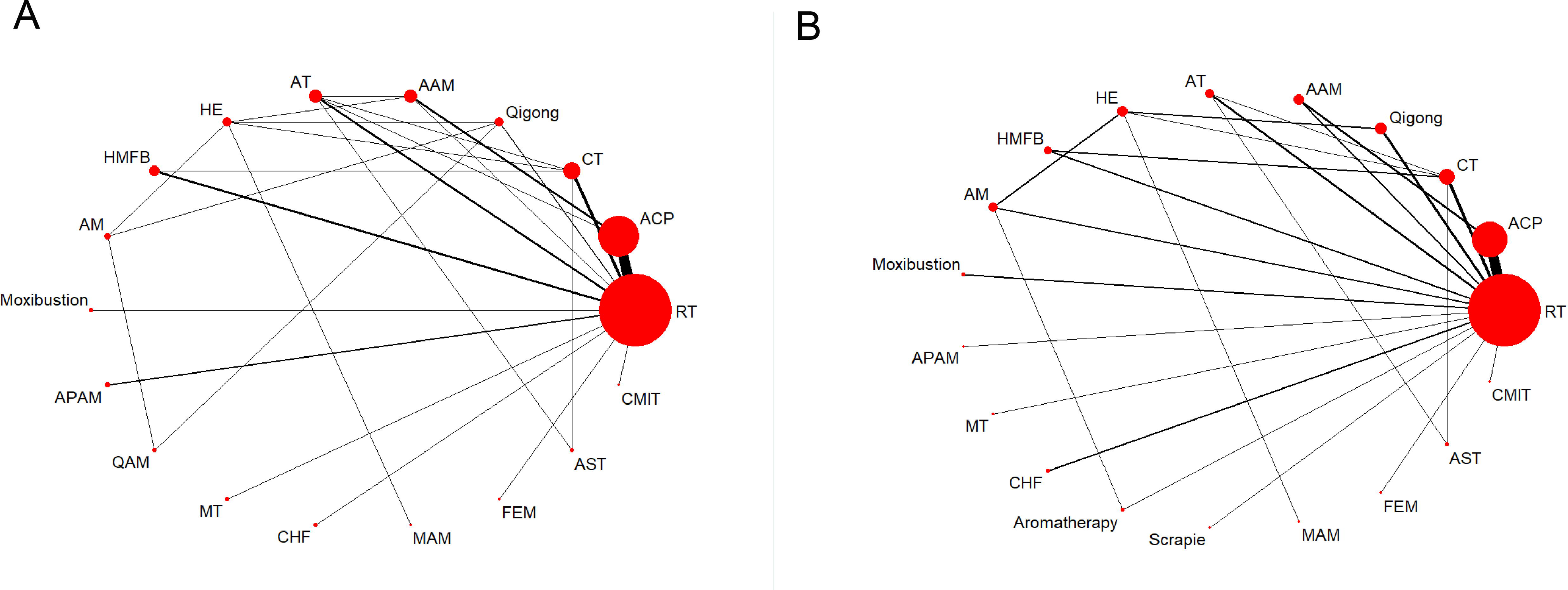
Network of eligible comparisons for TE rate (A) and PSQI scores (B). Line thickness represents trial numbers of treatment comparison, and the node size is proportional to the number of randomly assigned participants. Treatment abbreviation: Acupuncture (ACP), Combination therapy (CT), Acupuncture and moxibustion (AAM), Auricular therapy (AT), Health education (HE), Herbal medicine foot bath (HMFB), Acupressure massage (AM), Acupuncture plus acupressure massage (APAM), Qigong plus acupressure massage (QAM), Manipulative therapy (MT), Chinese herbal fumigation (CHF), Moxibustion plus acupressure massage (MAM), Five-element music (FEM), Acupoint sticking therapy (AST), Chinese medicine iontophoresis treatment (CMIT), patients who underwent TCM with routine treatment (RT). Treatments with full name: Moxibustion, Qigong, Aromatherapy, Scrapie.

As shown in Figure 3A, our network meta-analysis directly compared all the ETs-TCM with the control group. The results suggested that acupoint sticking therapy (OR = 0.24, 95%CI: 0.06-0.98), manipulative therapy (OR = 0.27, 95%CI: 0.08-0.92), acupressure massage (OR = 0.13, 95%CI: 0.03-0.50), auricular therapy (OR = 0.19, 95%CI: 0.10-0.40), acupuncture and moxibustion (OR = 0.14, 95%CI: 0.07-0.28), Qigong (OR = 0.13, 95%CI: 0.05-0.29), combination therapy (OR = 0.29, 95%CI: 0.16-0.52) and acupuncture (OR = 0.30, 95%CI: 0.21-0.44) were effective (Figure 4). In addition, acupuncture and moxibustion (OR = 0.15, 95%CI: 0.03-0.73) and Qigong (OR = 0.14, 95%CI: 0.03-0.71) were significantly more beneficial than Chinese herbal fumigation; Qigong plus acupressure massage was significantly more beneficial than acupuncture plus acupressure massage (OR = 22.66, 95%CI: 1.44-356.26), herbal medicine foot bath (OR = 27.70, 95%CI: 1.94-395.52), health education (OR = 49.59, 95%CI: 3.98-617.64), combination therapy (OR = 16.40, 95%CI: 1.24-217.38) and acupuncture (OR = 17.05, 95%CI: 1.31-222.68); acupressure massage (OR = 6.75, 95%CI: 2.24-20.32) was significantly more beneficial than health education; acupuncture and moxibustion (OR = 0.28, 95%CI: 0.10-0.81) and Qigong (OR = 0.26, 95%CI: 0.08-0.81)were significantly more beneficial than herbal medicine foot bath; auricular therapy (OR = 0.22, 95%CI: 0.07-0.75), acupuncture and moxibustion (OR = 0.16, 95%CI: 0.05-0.49), Qigong (OR = 0.14, 95%CI: 0.05-0.41) and combination therapy (OR = 0.33, 95%CI: 0.11-0.96) also were significantly more beneficial than health education.

**Figure 4:**
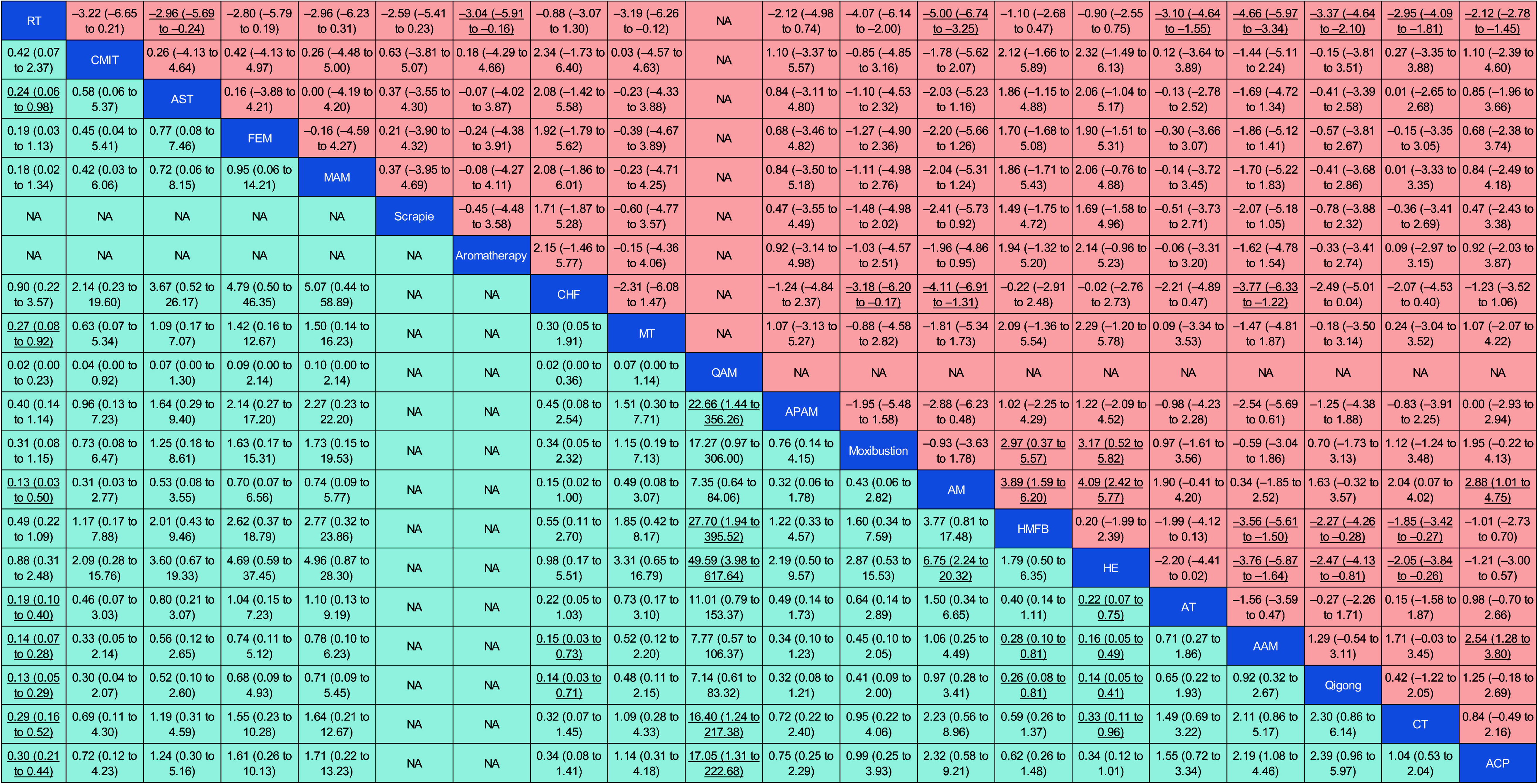
A network meta-analysis of TE rate and PSQI scores. Interventions are presented in alphabetical order. Treatment comparisons should be read from left to right, and the estimate intersects the column-defining treatment and the row-defining treatment. For the TE rate (blue), an OR less than 1 favors the row-defining treatment. For PSQI scores (red), the SMD less than 0 favours the column-defining treatment. Significant results are underlined.

The comparative effects among various ETs-TCM are presented in Figure 4. To further evaluate the relative effectiveness of these treatments, we also calculated the ranking of each ET-TCM based on cumulative probability plots and SUCRAs, which are presented in Figure 5A and Table 2. According to these calculations, Qigong plus acupressure massage had the highest probability (83.5%) of being the most effective ET-TCM for TE rate, with a SUCRA value of 96.7 (Table 2).

**Figure 5:**
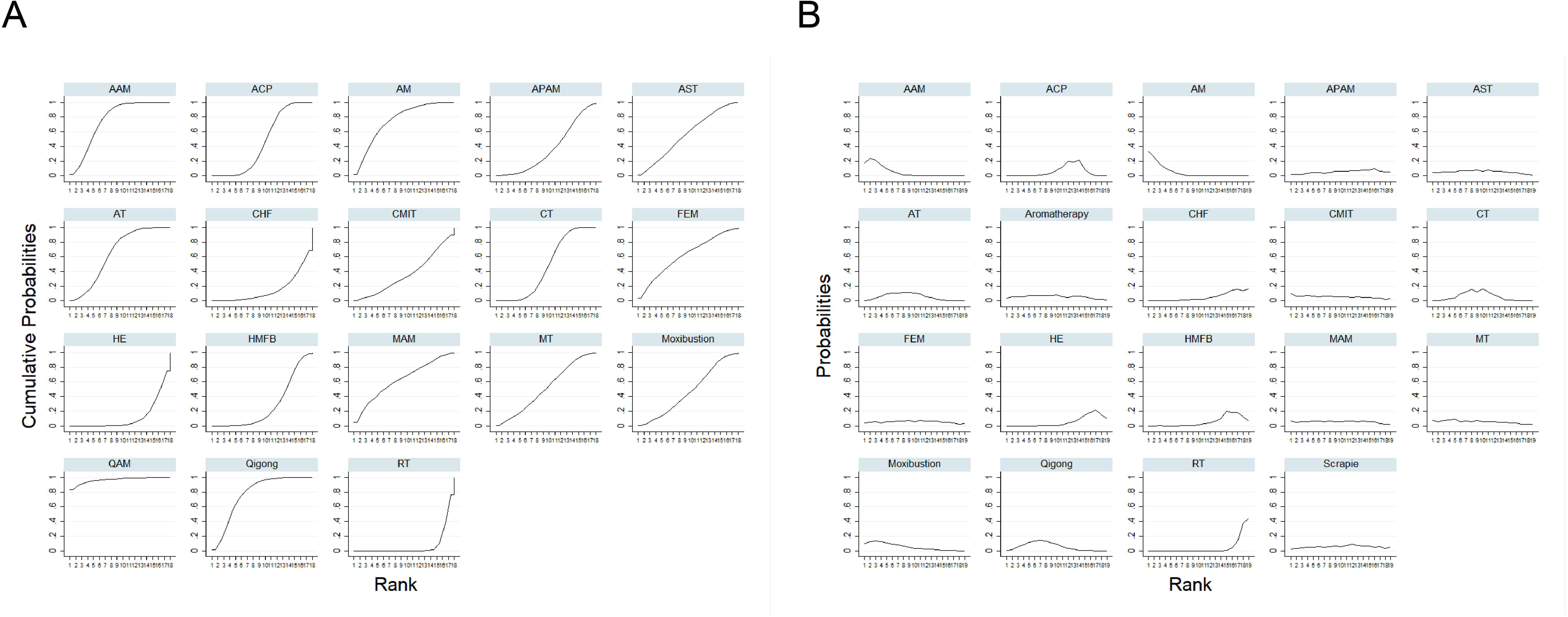
Cumulative ranking probability plots for TE rate (A) and PSQI scores(B). The horizontal axis represents the possible rank of each treatment (from best to worst according to the outcome). The vertical axis represents the cumulative probability of each treatment being the best option, the best of 2 options, the best of 3 options, and so on.

### 3.3. PSQI scores of ETs-TCM for insomnia in the elderly

For the analysis of PSQI scores, 60 studies with 4,524 participants were included. 58 studies adopted a 2-arm design, and two adopted a 3-arm design to compare various ETs-TCM (Appendix 3.2). The pairwise analysis is indicated in Appendix 3.2. An inconsistency test based on network analysis revealed no significant global inconsistency; the detailed results are shown in Appendix 4.2. Additionally, the funnel plot showed no evidence of publication bias (Appendix 5.2).

The network plot for PSQI scores (Figure 3B) showed all the available comparisons from the included trials. In Figure 3B, all the ETs-TCM were directly compared with the control group. And the results of our network meta-analysis suggested that acupoint sticking therapy (MD = –2.96, 95%CI:–5.69 to –0.24), Aromatherapy (MD = –3.04, 95%CI: (–5.91 to –0.16), acupressure massage (MD = –5.00, 95%CI: –6.74 to – 3.25), auricular therapy (MD = –3.10, 95%CI: –4.64 to –1.55), acupuncture and moxibustion (MD = –4.66, 95%CI: –5.97 to –3.34), Qigong (MD = –3.37, 95%CI: –4.64 to –2.10), combination therapy (MD = –2.95, 95%CI: –4.09 to –1.45) and acupuncture (MD = –2.12, 95%CI: –2.78 to –1.45) were effective (Figure 4).

In addition, moxibustion was significantly more beneficial than herbal medicine foot bath (MD = 2.97, 95%CI: 0.37 to 5.57) and health education (MD = 3.17, 95%CI: 0.52 to 5.82); acupressure massage was significantly more beneficial than herbal medicine foot bath (MD = 3.89, 95%CI: 1.59 to 6.20), health education (MD = 4.09, 95%CI: 2.42 to 5.77) and acupuncture (MD = 2.88, 95%CI: 1.01 to 4.75); acupuncture and moxibustion (MD = –3.56, 95%CI: –5.61 to –1.50), Qigong (MD =–2.27, 95%CI: – 4.26 to –0.28) and combination therapy (MD = –1.85, 95%CI: –3.42 to –0.27) also were significantly more beneficial than herbal medicine foot bath; acupuncture and moxibustion (MD = –3.76, 95%CI: –5.87 to –1.64), Qigong (MD =–2.47, 95%CI: –4.13 to –0.81) and combination therapy (MD = –2.05, 95%CI: –3.84 to –0.26) were significantly more beneficial than health education; acupuncture and moxibustion was significantly more beneficial than acupuncture (MD = 2.54, 95%CI: 1.28 to 3.80).

The comparative PSQI scores among various ETs-TCM are presented in Figure 4. The ranking of the ETs-TCM based on cumulative probability plots and SUCRAs are shown in Figure 5B and Table 2. According to the calculations, acupressure massage had the highest probability (32.9%) of being the most effective ET-TCM for reducing PSQI scores, with a SUCRA value of 90.9 (Table 2).

## 4. Discussion

Our network meta-analysis compared the efficacy of various ETs-TCM therapies on senile insomnia based on TE and PSQI. With 13,846 terms searched across 9 databases, we read 2,936 full-text articles and recruited 12,724 participants from 85 RCTs (46 for TE, 60 for PSQI) in our meta-analysis. Our study systematically compared 19 ETs-TCM therapies and found that Qigong plus acupressure massage (SUCRA=96.7) and Qigong (SUCRA=78) were the most effective ETs-TCM analyzed from the TE. Analyzed from PSQI, we also found acupressure massage (SUCRA=90.9) and “acupuncture and moxibustion” (SUCRA=87.4) were the top two effective ETs-TCM. Additionally, we highlight the benefits of manual therapy and gentle mind-body exercise as suitable options for treating insomnia in the elderly, such as the practice of health Qigong or Tai Chi.

Currently, a growing body of research is dedicated to exploring the effectiveness of various treatments for insomnia. While there is limited research focusing on the effectiveness of ETs-TCM therapies for senile insomnia. Previous studies have evaluated the effects of the western medicine [114], non-pharmacological treatment [115], or pharmacological and non-pharmacological interventions [116] for insomnia in the elderly. However, comparing multiple external TCM therapies has largely been overlooked, and no specific ETs-TCM therapy has been identified as the optimal treatment for senile insomnia. Our network meta-analysis aimed to address this gap by comparing and evaluating the optimal ETs-TCM for insomnia in the geriatric population. To include all available ETs-TCM therapy, we used a wide range of search terms. Our study searched a total of 13846 papers up to April 27, 2023, including 12,724 participants from 9 databases, providing a robust foundation for further data analysis and understanding of the effectiveness of ETs-TCM therapies for insomnia in the elderly. Also, we include two standards (TE, PSQI) to evaluate the ETs-TCM therapies on sleep quality, allowing us to include a greater number of therapies and evaluate them more comprehensively. To the best of our knowledge, this is the most comprehensive network meta-analysis focusing on external TCM therapies with such a large number of participants.

Compared to the reference treatment group (TE=7.4 and PSQI=4.3), all ETs-TCM therapies processed higher values in TE and PSQI, demonstrating their effectiveness in treating senile insomnia. While health education was found to be the least effective in improving sleep, either viewed from TE (12) or PSQI (16.4). Our study found that Qigong plus acupressure massage (SUCRA=96.7), Qigong (SUCRA=78.0), and acupuncture and moxibustion (SUCRA=76.6) were the best interventions when evaluated by the TE rate. Similarly, acupressure massage (SUCRA=90.9), acupuncture and moxibustion (SUCRA=87.4), and Qigong (SUCRA=64.7) were the top priorities interventions when evaluated by the PSQI score. This suggests that both physical stimulation and gentle mind-body exercise, as provided by acupressure massage and Qigong, may positively affect sleep quality. It is possible that the geriatric population may not fully understand or adhere to the health education, highlighting the importance of tailoring treatments to the unique characteristics of the target population.

We also found that acupressure massage ranks top in PSQI but fourth in TE, with less efficacy than Qigong plus acupressure massage. This raises the question of whether therapy combination promotes the effects as a general rule. We therefore further classify 19 included ETs-TCM therapies into monotherapy and combination therapy. The monotherapy included acupuncture, Qigong, auricular therapy, health education, herbal medicine foot bath, acupressure massage, moxibustion, manipulative therapy, Chinese herbal fumigation, aromatherapy, scrapie, five-element music, acupoint sticking therapy, and Chinese medicine iontophoresis treatment. The remaining combination therapy included “acupuncture and moxibustion”, “acupuncture plus acupressure massage”, “Qigong plus acupressure massage”, and “moxibustion plus acupressure massage”. Among them, Qigong plus acupressure massage was only included in TE (n=17); Aromatherapy and Scrapie were only included in PSQI (n=18). We found that not all combination therapy may be more effective based on the ranking of SUCRA. Some combination therapy did have sound effects; for example, acupuncture and moxibustion (TE:3/17, PSQI:2/18) = acupuncture (TE:12/17, PSQI:15/18) + moxibustion (TE:11/17, PSQI:3/18). However, some combinations were not as effective: such as moxibustion plus acupressure massage (TE:6/17, PSQI:10/18) = Moxibustion (TE:11/17, PSQI:3/18) + acupressure massage (TE:4/17, PSQI: 1/18) and acupuncture plus acupressure massage (TE:13/17, PSQI:14/18) = acupuncture (TE:12/17, PSQI:15/18) + AM (TE:4/17, PSQI: 1/18), which were not as effective as acupressure massage alone. The quality of the included studies, such as sample size, study design, and potential biases, may explain some discrepancies in the effectiveness of combination therapies. Additionally, the variation in TCM differential diagnosis for elderly patients, which is influenced by individual patient characteristics, may also contribute to these discrepancies.

Qigong+ acupressure massage (TE:1/17) or Qigong (TE:2/17, PSQI:4/18) have been firmly proven effective, highlighting the significance of Qigong itself. Health Qigong is a traditional Chinese practice that involves gentle movements, breathing techniques, and meditation to promote physical, mental, and spiritual health [117, 118]. It encompasses various styles, including Tai Chi, Ba Duan Jin, Liuzijue, Wuqinxi, etc [119]. These gentle mind-body exercises may be particularly suitable for treating insomnia in the elderly. They have been shown to improve sleep quality, reduce anxiety and depression, and increase overall well-being [120, 121]. Health Qigong’s ability to improve sleep quality has been attributed to its ability to regulate the autonomic nervous system [122] and activate the parasympathetic nervous system [123], reducing stress and anxiety. Additionally, the slow, repetitive movements and deep breathing exercises performed in health Qigong have improved sleep onset and duration by promoting physical relaxation and reducing mental tension [124]. In this sense, health Qigong can be considered a form of mindfulness-based intervention. Its integration of physical, mental, and spiritual elements may provide a unique and effective approach to treating insomnia in the elderly. As a safe and low-risk form of exercise, health Qigong may be a particularly appealing option for those seeking to improve their sleep quality without medication.

Despite our study’s significant findings, several limitations must be addressed. Firstly, the sample size of different treatments varied a lot, with some treatments having limited sample sizes, such as Scrapie and “moxibustion plus acupressure massage”, which may limit the generalizability of our results. Secondly, some external treatments were only evaluated using one standard, such as Qigong plus acupressure massage, which was only evaluated using the TE. This raises questions about the superiority of these simple-evaluated treatments in other uninvolved standards. Thirdly, our study utilized two standards for evaluating the efficacy of external TCM therapies but needed a comprehensive method for reconciling potential discrepancies between the two standards. This may lead to uncertainty in the results when conflicts arise between the two metrics of TE and PSQI. Additionally, while our study provides insights into the efficacy of various external TCM therapies, we acknowledge that clinical practice may differ. The effectiveness of treatments can vary depending on patient characteristics and local practices, meaning our findings may not fully reflect real-world clinical outcomes. Lastly, the mechanisms of many ET-TCM therapies remain largely unknown, and future research should focus on uncovering these mechanisms to further understand their effectiveness. Future studies could focus on increasing the sample size and incorporating a more diverse population to address these limitations. To further explore the effects of health Qigong on insomnia, it would be valuable to compare different types of health Qigong and investigate their relative efficacy. Randomized controlled trials with larger sample sizes and more extended follow-up periods should be conducted further to confirm the effectiveness of these external TCM therapies on the older. In this way, we may provide more concrete, evidence-based recommendations for treating insomnia in the elderly, promoting better health outcomes for this vulnerable group.

## 5. Conclusion

Our study demonstrates that external TCM therapies, specifically Qigong plus acupressure massage, Qigong, acupuncture and moxibustion, can effectively reduce insomnia symptoms in the elderly. Moreover, our findings suggest that monotherapy may be more effective than combination therapy, with Qigong being a suitable option for older adults. This highlights the importance of incorporating manual therapy and gentle mind-body exercise as alternative solutions to medication, particularly for elderly individuals susceptible to drug side effects and misuse.

## Author contributions

Xinxin Ye undertook the study design, completed literature searching, data analysis, drafted and revised this paper. Huanju Liu completed literature searching, data extraction, drafted and revised this paper. Zhuzhu Qin, Yining Tao, Danfeng Chen, Jiaxin Zhang, Xiaofei Liang, Hui Li, Ruizhe Jiang, and Ping Wang completed literature searching, data extraction and revised this paper. Cong Huang undertook the study design, data analysis and revised this paper.

## Financial support

This study was supported by the fundamental research funds (204201∗172220192) for the Central Universities, China, and the Hundred Talents Program funding (188020∗194221802/004/001) from Zhejiang University, China.

## Conflicts of interest

The authors declare no conflicts of interest in this study.

## Acknowledgments

The authors wish to acknowledge the support of Zhejiang University.

## Data availability

Any additional information can be obtained from the author on request.

## Notes

### Competing Interest Statement

The authors have declared no competing interest.

## Reference

[1] Z. Alimoradi, E. Jafari, A. Brostrom, M.M. Ohayon, C.Y. Lin, M.D. Griffiths, K. Blom, S. Jernelov, V. Kaldo, A.H. Pakpour, Effects of cognitive behavioral therapy for insomnia (CBT-I) on quality of life: A systematic review and meta-analysis, Sleep Med Rev, 64 (2022) 101646.

[2] D.J. Buysse, Insomnia, Jama, 309(7) (2013) 706–16.

[3] C.M. Morin, R. Benca, Chronic insomnia, Lancet (London, England), 379(9821) (2012) 1129-41.

[4] K. Crowley, Sleep and sleep disorders in older adults, Neuropsychol Rev, 21(1) (2011) 41–53.

[5] A. Hombali, E. Seow, Q. Yuan, S.H.S. Chang, P. Satghare, S. Kumar, S.K. Verma, Y.M. Mok, S.A. Chong, M. Subramaniam, Prevalence and correlates of sleep disorder symptoms in psychiatric disorders, Psychiatry Res, 279 (2019) 116–122.

[6] B. Zheng, C. Yu, J. Lv, Y. Guo, Z. Bian, M. Zhou, L. Yang, Y. Chen, X. Li, J. Zou, F. Ning, J. Chen, Z. Chen, L. Li, Insomnia symptoms and risk of cardiovascular diseases among 0.5 million adults, Neurology, 93(23) (2019) e2110.

[7] Y. Wang, T. Jiang, X. Wang, J. Zhao, J. Kang, M. Chen, H. Wang, L. Niu, Y. Wang, Y. Zhou, J. Wu, H. Fu, Z. Cai, Z. Li, J. Chen, Association between Insomnia and Metabolic Syndrome in a Chinese Han Population: A Cross-sectional Study, Sci Rep, 7(1) (2017) 10893.

[8] J.M. Trauer, M.Y. Qian, J.S. Doyle, S.M. Rajaratnam, D. Cunnington, Cognitive Behavioral Therapy for Chronic Insomnia: A Systematic Review and Meta-analysis, Ann Intern Med, 163(3) (2015) 191–204.

[9] S. Madari, R. Golebiowski, M.P. Mansukhani, B.P. Kolla, Pharmacological Management of Insomnia, Neurotherapeutics, 18(1) (2021) 44–52.

[10] L.F. Maurer, J. Schneider, C.B. Miller, C.A. Espie, S.D. Kyle, The clinical effects of sleep restriction therapy for insomnia: A meta-analysis of randomised controlled trials, Sleep Med Rev, 58 (2021) 101493.

[11] J. Wang, Y. Chen, X. Zhai, Y. Chu, X. Liu, X. Ma, Visualizing Research Trends and Identifying Hotspots of Traditional Chinese Medicine (TCM) Nursing Technology for Insomnia: A 18-Years Bibliometric Analysis of Web of Science Core Collection, Front Neurol, 13 (2022) 816031.

[12] Z. Dai, X. Liao, L.S. Wieland, J. Hu, Y. Wang, T.H. Kim, J.P. Liu, S. Zhan, N. Robinson, Cochrane systematic reviews on traditional Chinese medicine: What matters-the quantity or quality of evidence?, Phytomedicine, 98 (2022) 153921.

[13] Q. Wu, X. Chen, G. Gan, Q. Zhang, L. Yu, C. Li, Y. Zhang, M. Ao, Visual analysis and evaluation of clinical research on Traditional Chinese medicine compounds in treating insomnia of Yin deficiency syndrome, J Ethnopharmacol, 298 (2022) 115669.

[14] X. Luan, X. Zhang, Y. Zhou, The Role and Clinical Observation of Traditional Chinese Medicine in Relieving Senile Insomnia: A Systematic Review and Meta-Analysis, Biomed Res Int, 2022 (2022) 9484095.

[15] X. Liu, Y. Wang, Q. Ye, Y. Sun, J. Yang, Y. Dai, Q. Wen, External treatment of traditional Chinese medicine for constipation after thoracolumbar compression fractures: A protocol for systematic review and meta-analysis, Medicine (Baltimore), 100(35) (2021) e27110.

[16] X. Yin, M. Gou, J. Xu, B. Dong, P. Yin, F. Masquelin, J. Wu, L. Lao, S. Xu, Efficacy and safety of acupuncture treatment on primary insomnia: a randomized controlled trial, Sleep Med, 37 (2017) 193–200.

[17] H. Feng, A. Pan, G. Zheng, W. Yu, Clinical study of auricular point seed burying combined with fire dragon pot moxibustion in perimenopausal women with insomnia, J Obstet Gynaecol Res, 48(7) (2022) 1938–1944.

[18] L.M. Deuel, L.C. Seeberger, Complementary Therapies in Parkinson Disease: a Review of Acupuncture, Tai Chi, Qi Gong, Yoga, and Cannabis, Neurotherapeutics, 17(4) (2020) 1434–1455.

[19] W. Li, Y. Cheng, Y. Zhang, Y. Qian, M. Wu, W. Huang, N. Yang, Y. Liu, Shumian Capsule Improves the Sleep Disorder and Mental Symptoms Through Melatonin Receptors in Sleep-Deprived Mice, Front Pharmacol, 13 (2022) 925828.

[20] Y. Sun, N. Zhang, Y. Qu, Y. Cao, J. Li, Y. Yang, T. Yang, Y. Sun, Shuangxia decoction alleviates p-chlorophenylalanine induced insomnia through the modification of serotonergic and immune system, Metab Brain Dis, 35(2) (2020) 315–325.

[21] B. Rouse, A. Chaimani, T. Li, Network meta-analysis: an introduction for clinicians, Intern Emerg Med, 12(1) (2017) 103–111.

[22] J.P. Higgins, D.G. Altman, P.C. Gøtzsche, P. Jüni, D. Moher, A.D. Oxman, J. Savovic, K.F. Schulz, L. Weeks, J.A. Sterne, The Cochrane Collaboration’s tool for assessing risk of bias in randomised trials, Bmj, 343 (2011) d5928.

[23] D.J. Buysse, C.F. Reynolds, 3rd, T.H. Monk, S.R. Berman, D.J. Kupfer, The Pittsburgh Sleep Quality Index: a new instrument for psychiatric practice and research, Psychiatry Res, 28(2) (1989) 193–213.

[24] B. Dong, H. Li, W. Gao, Z. Zhou, The effect of intra-articular injection of antibacterial drugs in the clinical treatment of prosthetic joint infection in patients undergoing artificial hip replacement, Am J Transl Res, 13(4) (2021) 3508–3514.

[25] T.A. Furukawa, C. Barbui, A. Cipriani, P. Brambilla, N. Watanabe, Imputing missing standard deviations in meta-analyses can provide accurate results, J Clin Epidemiol, 59(1) (2006) 7–10.

[26] S. Shim, B.H. Yoon, I.S. Shin, J.M. Bae, Network meta-analysis: application and practice using Stata, Epidemiol Health, 39 (2017) e2017047.

[27] J.P. Higgins, D. Jackson, J.K. Barrett, G. Lu, A.E. Ades, I.R. White, Consistency and inconsistency in network meta-analysis: concepts and models for multi-arm studies, Res Synth Methods, 3(2) (2012) 98–110.

[28] G. Salanti, A.E. Ades, J.P. Ioannidis, Graphical methods and numerical summaries for presenting results from multiple-treatment meta-analysis: an overview and tutorial, J Clin Epidemiol, 64(2) (2011) 163–71.

[29] X.B. Wang, H.G. Huang, Y.J. Yang, Efficacy of foot bath with TCM medicine on elderly patients with insomnia, Clinical Journal of Chinese Medicine, 13(07) (2021) 95–97.

[30] Y.L. Wang, R. Zhang, C.X. Wang, H.G. Hu, M.J. Xiao, R.Q. Sui, Clinical observation of acupuncture at Dongshiqi point combined with Sini powder in the treatment of senile insomnia, Chinese Journal of Geriatric Care, 19(03) (2021) 17–19.

[31] Y. Wang, J.L. Bao, J.H. Hu, M.X. Wang, Q. Yan, X.M. Zhang, Effect of pressure inoculation training combined with auricular point pressing bean in elderly patients with diabetes and insomnia Chinese Journal of Gerontology, 41(5) (2021) 955–958.

[32] X.F. Lin, Y. Qing, J. Zhong, Clinical observation of modified liuwei dihuang pill combined with electroacupuncture in the treatment of senile hypertension complicated with insomnia, Chinese Community Doctors, 37(13) (2021) 92–93+96.

[33] S.D. Li, The clinical efficacy observation of tiaoqi guiyuan prescription Acupuncture in the treatment on senile insomnia with incoordination between heart and kidney, Fujian University of Traditional Chinese Medicine, (_issue %/ _ori_publication) (2021).

[34] P.M. Siu, A.P. Yu, B.T. Tam, E.C. Chin, D.S. Yu, K.-F. Chung, S.S. Hui, J. Woo, D.Y. Fong, P.H. Lee, G.X. Wei, M.R. Irwin, Effects of Tai Chi or Exercise on Sleep in Older Adults With Insomnia A Randomized Clinical Trial, JAMA NETWORK OPEN, 4(e20371992) (2021).

[35] J. Wang, Y. Liu, Y.L. Zhang, N. Zhuang, Effect of acupuncture and moxibustion combined with psychological counseling on insomnia in the aged, Chinese Journal of Gerontology, 40(12) (2020) 2598–2600.

[36] Q.C. Gao, Clinical research on the effects of acupuncture on sleep quality and cognitive function for senile primary insomnia, Heilongjiang University Of Chinese Medicine, (_issue %/ _ori_publication) (2020).

[37] Y. Zheng, Y.J. Tan, Q.P. Wei, D.J. Wang, L. Chen, Clinical observation of Zhao’s thunder fire moxibustion in the intervention of senile insomnia with deficiency of heart and spleen, Journal of Guangxi University of Chinese Medicine, 23(02) (2020) 20–22.

[38] S. Cai, Clinical study on cupreous scraping therapy for insomnia in the elderly, Guangzhou University of Chinese Medicine, (_issue %/ _ori_publication) (2020).

[39] Y.J. Ding, Study on the intervention effect of Taijiquan on insomnia of the elderly, Anhui University of Chinese Medicine, (_issue %/ _ori_publication) (2020).

[40] L.L. Zhu, Clinical effect of cinnamon powder applied to Yongquan point combined with auricular point on senile insomnia, Chinese Community Doctors, 36(34) (2020) 111–112.

[41] F.B. Yan, H. Chen, Y. Zhu, H.Y. Shi, W.Y. Cheng, R.M. Bian, The application of the appropriate technical serv ice model of traditional chinese medicine in the treatment of sleep disorder under the family doctor system, Chinese Medicine Modern Distance Education of China, 18(15) (2020) 71–73.

[42] Y.Q. Lu, Clinical study of acupuncture combined with ear acupuncture on sleep disorders in patients with Alzheimer’s disease based on dumai theory, Heilongjiang University Of Chinese Medicine, (_issue %/ _ori_publication) (2020).

[43] X. Wang, X.N. Cui, L.L. Shang, L.H. Zheng, J. Chen, M.L. Tian, R.Q. Guan, F. Sun, Clinical observation of auricular acupoint pressing combined with meridian flow therapy for treating senile insomnia with heart-kidney disjunction, China’s Naturopathy, 28(24) (2020) 56–58.

[44] B.L. Sun, J.L. Bao, L. Wang, R. Yin, Y. Wang, G.H. Xiong, Effects of Baihui acupoint (GV20) moxibustion combined with acupoint massage on anxiety and sleeping quality of elderly female patients with insomnia, Journal of Guangzhou University of Traditional Chinese Medicine, 37(04) (2020) 676–680.

[45] Z.H. Song, Y.J. Zhang, X.H. Liu, Clinical study of "Introducing Yang into Yin" massage combined with five-tone therapy on middle-aged and elderly patients with hypertension and insomnia Proceedings of the 2020 Annual Academic Conference of the Gansu Academy of Traditional Chinese Medicine, (_issue) (2020) 6 %/ _ori_publication.

[46] H.S. Zhang, P. Liu, D.Y. Cong, Clinical observation on the curative effect of abdominal massage with abdominal vibration and ring kneading in the treatment of primary insomnia with deficiency of heart and spleen in middle-aged and old people, Chinese Journal of Gerontology, 39(19) (2019) 4773–4775.

[47] Q.S. He, Z.Y. Du, Clinical observation on treating middle-aged insomnia by acupuncture plus cupping, Clinical Journal of Chinese Medicine, 11(28) (2019) 117–119.

[48] X.P. Yu, Q.C. Gao, Clinical study on the effect of acupuncture on sleep quality and cognitive function in senile patients with primary insomnia, Jiangsu Journal of Traditional Chinese Medicine, 51(04) (2019) 62–64.

[49] J.J. Hou, Y. Bai, Y.E. Lu, Y. Lv, Q.Y. Yang, R.W. Xue, B.Q. Zhi, Effect of yuchai fushen granule combined with sanbu acupoint selection acupuncture on cerebral hemodynamic is and quality of life in elderly patients with chronic insomnia, Acta Chinese Medicine, (_issue) (2019) 5 %/ _ori_publication.

[50] J.Z. Fan, J.M. Xiong, X. He, Clinical Study on Treatment of Senile Insomnia with Yangxin Hewei Yimian Decoction Combined with Acupuncture at Hypnotic Acupoint, Chinese Archives of Traditional Chinese Medicine, 37(2) (2019) 466–469.

[51] Y.L. Liu, X.S. Liu, Effect of five elements music therapy combined with Chinese materia medica foot bath method on sleep quality of elderly people in community, Journal of Changchun University of Chinese Medicine, 35 (2019) 338–340.

[52] Y.W. Luo, Clinical observation of warm acupuncture and moxibustion in treating senile insomnia of Yang Deficiency type, China’s Naturopathy, 27(18) (2019) 22–24.

[53] Z.C. Ye, Clinical effect analysis of the treatment of primary insomnia by screw acupuncture combined with massage, Chinese Manipulation and Rehabilitation Medicine, 10(02) (2019) 17–18.

[54] Y.M. Chen, Y.Y. Qu, Z.M. Qu, Analysis on the curative effect of Qijudihuang Decoction plus and minus foot massage in treating senile hypertension with insomnia, Chinese Journal of Modern Drug Application, 13(13) (2019) 132–133.

[55] C.M. Pan, F.H. Zeng, W.X. Zhou, A.J. Jin, Clinical observation of Modified Huanglian Decoction combined with acupuncture and moxibustion in the treatment of middle-aged and elderly women with insomnia, Clinical Journal of Chinese Medicine, 11(24) (2019) 101–103.

[56] J. Chen, L.K. Zhou, D. Zhang, H. Zhao, X.R. Gao, Dong shi qi point with nourishing heart and tranquilizing expectorant soup clinical effect analysis of intervention on insomnia of phlegm-dampness in the aged, Journal of Hebei Traditional Chinese Medicine and Pharmacology, 34(04) (2019) 37–40.

[57] D.L. Wei, Research on the intervention of Baduanjin combined with cognitive behavioral therapy in insomnia of the elderly, Journal of Beijing University of Traditional Chinese Medicine, (_issue %/ _ori_publication) (2019).

[58] Y.W. Wang, X.D. Li, X.J. Bi, Effects of Baduanjin fitness Qigong on sleep quality and memory function of elderly people in community, Chinese Journal of Gerontology, 39 (2019) 3435–3437.

[59] X.Y. You, X.K. Chen, C.Y. Wang, Clinical observation on the treatment of insomnia in elderly patients by acupuncture method of “Xingnao Kaiqiao Pairing Points”, World Latest Medicine Information, 19(78) (2019) 144–145+151.

[60] Z.Z. Jia, J.X. Liu, The application effect of traditional Chinese medicine foot bath combined with traditional Chinese medicine internal medicine nursing in senile insomnia, China Continuing Medical Education, 10(14) (2018) 165–166.

[61] Q.L. Liao, Observation on treating elderly insomnia with the tianwang buxin decoction plus acupuncture, Clinical Journal of Chinese Medicine, 10(20) (2018) 65–67.

[62] F.Z. Gao, Study on the effect of practicing Baduanjin combined with acupoint massage therapy on sleep quality of middle-aged and elderly patients with insomnia in community, Journal of Beijing University of Traditional Chinese Medicine, (_issue %/ _ori_publication) (2018).

[63] H. Zhao, L.M. Guo, Q. Zhang, Nursing analysis of moxa stick moxibustion combined with auricular point pressure bean treatment on insomnia with deficiency syndrome of elderly patients with coronary heart disease, Clinical Journal of Chinese Medicine, 10(36) (2018) 73–74.

[64] W.H. Pang, C. Wei, Treating 39 cases of senile insomnia with sisheng yin and auricular point sticking, Clinical Journal of Chinese Medicine, 10(05) (2018) 72–73.

[65] F. Yuan, X.Y. Zhao, Y.L. Huang, B. Luo, Clinical evaluation on guipi decoction combined with acupuncture in the treatment of elderly patients with insomnia(syndrome of deficiency of both qi and blood), China Pharmaceuticals, 27(19) (2018) 34–37.

[66] J.M. Li, S. Zhou, F. Liu, H.H. Luo, J.Y. Luo, Clinical observation and nursing experience of treating senile sleep disorder with acupuncture and moxibustion, Yunnan Journal of Traditional Chinese Medicine and Materia Medica, 39(11) (2018) 92–94.

[67] X.H. Miao, Efficacy of baduan Jin and wuxing music therapy on insomnia in elderly hypertension, Clinical Journal of Chinese Medicine, 10(31) (2018) 136–138.

[68] Y.T. Tan, Observation on application effect of Chinese medicine iontophoresis for elderly patients with insomnia after stroke, Chinese Nursing Research, 31(22) (2017) 2737–2740.

[69] Y. Wang, Clinical Observation on Acupuncture and Medicine in the Treatment of Senile Coronary Heart Disease with Insomnia Syndrome, Chinese Medicine Modern Distance Education of China, 15(15) (2017) 98–100.

[70] H.J. Nie, Nursing intervention of traditional chinese medicine for the treatment of olderly patients with insomnia during perioperative period, Journal of Liaoning University of Traditional Chinese Medicine, 19(05) (2017) 219–221.

[71] W.X. Xue, J.Y. Zhang, L.L. Ge, Effect of acupuncture on sleep quality of senile neurotic insomnia, Chinese Journal of Gerontology, 37(21) (2017) 5390–5392.

[72] G.F. Li, The Clinical Study of Herbs-partition Moxibustion in treating Insomnia of deficiency of both the heart and spleen syndrome, Chengdu University of Traditional Chinese Medicine, (_issue %/ _ori_publication) (2017).

[73] X.M. Liang, Clinical observations on the therapeutic effect of ear acupoint thumbtack needle embedding on senile primary insomnia, Shanghai Journal of Acupuncture and Moxibustion, 36 (2017) 719–722.

[74] X.L. Yu, H.H. Li, The effect of Baduanjin combined with acupoint massage on the sleep quality in elderly patients with coronary heart disease, Practical Journal of Clinical Medicine, 14 (2017) 220–222.

[75] Y. Zhang, Clinical study of combined therapy of herbal fumigation and estazolam in the treatment of insomnia in the elderly, 康复医学与理疗学, Bengbu Medical College, 2016.

[76] W.J. Zhang, Analysis of curative effect of traditional Chinese medicine fumigation combined with cognitive behavioral therapy on senile insomnia, Journal of Practical Traditional Chinese Medicine, 32(4) (2016) 297–298.

[77] Z.J. Mou, N. Hui, T. Ma, Z.J. Kang, Fifty-one cases of insomnia in middle-aged and elderly patients were treated by acupuncture of Five zang Shu and Diaphragm Shu points plus Moxa box moxibustion, Yunnan Journal of Traditional Chinese Medicine and Materia Medica, 37(03) (2016) 52–53.

[78] X.X. Lin, The effect of listening to jiao tune on sleep quality of liver-fire type 2 diabetes elderly with insomnia, Fujian University of Traditional Chinese Medicine, (_issue %/ _ori_publication) (2016).

[79] F.F. Wang, P.D. Bian, X.F. Chen, Q.H. Miao, Rehabilitation treatment of senile patients with insomnia, Zhejiang Clinical Medical Journal, 18 (2016) 693–694.

[80] H. Li, L. Lin, Z. Li Y, M. Xiong, P. Tang, Efficacy of aromatherapy in improving elderly’s sleep quality, Journal of Chengdu Medical College, 11(01) (2016) 112–115.

[81] C.J. Zhang, L. Chang, Y.H. Sun, The clinical research on senile insomnia treated with integrative therapy of foot bath with chinese medicine and massage of foot points, Henan Traditional Chinese Medicine, 35(1) (2015) 210–212.

[82] J.P. Wang, J.B. Wang, L.C. Wang, Y.M. Zhang, Senile insomnia treated with integrated acupuncture and medication therapy: a randomized con trolled trial, Chinese Acupuncture & Moxibustion, 35(6) (2015) 544–548.

[83] Y.F. Gao, Y.P. Wu, Observation on 65 elderly hospitalized patients with insomnia treated by acupoint massage combined with Chinese medicine foot bath, Zhejiang Journal of Traditional Chinese Medicine, 50(07) (2015) 530–531.

[84] X.D. Luo, Observation on the curative effect of foot bath massage combined with auricular point sticking in the treatment of senile insomnia, Chinese Journal of Traditional Medical Science and Technology, 21 (2014) 436.

[85] J. He, D.Z. Zeng, G.Y. Gu, Y.L. Hu, J.W. Luo, Observation on curative effect of acupuncture therapy in the treatment of refractory insomnia in middle and old age, Modern Journal of Integrated Traditional Chinese and Western Medicine, 22(9) (2013) 957–959.

[86] Y. Li, J. Li, Observation on the curative effect of treating senile insomnia by syndrome differentiation and treatment combined with ear point burying bean, Journal of Basic Chinese Medicine, 18(10) (2012) 1126–1127.

[87] X.Y. Li, Z.L. Zhou, B. Zhang, Clinical observation on acupuncture for treating middle or elderly aged patients with insomnia, Tianjin Journal of Traditional Chinese Medicine, 27(5) (2010) 386–388.

[88] Z.Y. Sun, Acupuncture at taixi sanyinjiao yongquan point treated 40 cases of insomnia in the elderly, Shanxi Journal of Traditional Chinese Medicine, (6) (2010) 731–732.

[89] H. Chang, J.H. Wang, Y.P. Bai, Clinical observation on 144 cases of senile insomnia with deficiency of qi and blood treated by acupuncture and medicine, Hebei Journal of Traditional Chinese Medicine, 31 (2009) 673–674.

[90] S.L. Ye, Treatment of 58 cases of senile refractory insomnia by combination of acupuncture and medicine, Journal of Clinical Acupuncture and Moxibustion, 23 (2007) 16–17.

[91] L.Z. Song, H. Li, H. Zhang, X.R. Zhang, Acupuncture combined with relaxation therapy to treat sleep disorders in the elderly, Liaoning Journal of Traditional Chinese Medicine, 32 (2005) 359–360.

[92] H.L. Shan, D.M. Li, W.F. Liu, L. Li, H.L. Lin, The traditional Chinese medicine treatment of senile insomnia syndrome differentiation, Military Nursing, 19 (2002) 31–32.

[93] T. Han, F. Zhao, Treatment of 189 cases of senile insomnia by electroacupuncture plus ear pressure, Lishizhen Medicine and Materia Medica Research, 12 (2001) 446–447.

[94] X.L. Tan, J.L. Zhang, J.L. Cong, Y. Gao, J. Zhang, J. Cong, Y. Gao, Effect of drug-spate interposed moxibustion on Du channel on insomnia in the elderly, Journal of Nursing Science, 25 (2010) 60–61.

[95] C. Li, Y. Yu, F.L. Xu, Y. Yu, F. Xu, Observation on the curative effect of simple Taijiquan on insomnia of elderly people in community, Shanghai Medical & Pharmaceutical Journal, 31(S1) (2010) 66–67.

[96] W.J. Huang, Observation on curative effect of Chinese medicine foot bath on insomnia patients, Chinese Clinical Nursing, 1(01) (2009) 38–39.

[97] H.B. Zheng, Y. Chen, Y. Chen, Clinical observation of 241 cases of senile insomnia treated by electroacupuncture, Asia-Pacific Traditional Medicine, (09) (2008) 47–48.

[98] X.Y. Li, Clinical observation on acupuncture for treating middle or elderly aged patients with insomnia, Beijing University of Chinese Medicine, 2008.

[99] H. Jiang, Observation on curative effect of 89 cases of insomnia after cerebral apoplexy treated by massage manipulation and Chinese medicine Yunnan Journal of Traditional Chinese Medicine and Materia Medica, (04) (2004) 26–27.

[100] F.X. Long, S.J. Huang, X. Li, J. Lu, Q.F. Chen, S. Huang, X. Li, J. Lu, Q. Chen, Effect of auricular acupoint pressing bean and psychological nursing in community elderly patients with insomnia Clinical Nursing Research, 30(11) (2021) 64–65.

[101] L. Dong, Auricular seed embedding combined with acupoint massage can improve insomnia in elderly patients, Ease Up, (02) (2022) 110–111.

[102] L. Gui, C.F. Zhu, X.Y. Chen, Z.H. Xia, Q. Zhong, F. Guo, C. Zhu, X. Chen, Z. Xia, Q. Zhong, F. Guo, Clinical observation of umbilical abdomen moxibustion combined with acupuncture therapy on senile insomnia, Hubei Journal of Traditional Chinese Medicine, 43(01) (2021) 49–51.

[103] J.C. Hu, J.C. Hu, J. Hu, An empirical study on the intervention of Taijiquan in elderly patients with insomnia of heart and kidney disconnection, Sports Vision, (23) (2021) 71–73.

[104] F.F. Wang, Effect of tianmen combined with five-tone therapy nursing mode on sleep and quality of life of elderly patients with insomnia, Diet Health, (4) (2022) 69–72.

[105] D.M. Yang, Y.Z. Li, Y. Li, Clinical effect analysis of warm acupuncture and moxibustion on senile insomnia patients with Yang deficiency, Health Guide, (46) (2021) 113–114.

[106] X.Y. Li, Clinical study on acupuncture treatment of insomnia in middle and old age, Healthmust-Readmagazine, (11) (2021) 10.

[107] L. Ding, X.Y. Liu, S.Q. Dai, X.S. Qu, Y. Han, L. Xue-yan, D. Shu-qing, Q.U. Xian-shuang, H. Ye, Observation on the Effect of Acupuncture and Medicine Combined Treatment of Senile Essential Hypertension with Insomnia, World Journal of Integrated Traditional and Western Medicine, 17(1) (2022) 137–141,146.

[108] T.T. Hu, B. Han, B. Han, Effect analysis of traditional Chinese medicine foot bath on senile patients with sleep disorder, Kang Yi, (3) (2021) 230.

[109] X. Wen, W. Huang, X.D. Han, W. Huang, X. Han, Observation on the curative effect of treating senile insomnia by TCM syndrome differentiation and treatment combined with ear point burying bean, Kang Yi, (6) (2021) 204.

[110] G.L. Wu, W.D. Pan, H.L. Yu, Y.F. He, W. Pan, H. Yu, Y. He, Analysis of the therapeutic effect of "Jin Sanzhen" Tiaoshen Acupuncture on sleep disorders in elderly patients with post-stroke depression, Clinical Journal of Chinese Medicine, 13(21) (2021) 95–97.

[111] B. Fan, W. Song, J. Zhang, Y. Er, B. Xie, H. Zhang, Y. Liao, C. Wang, X. Hu, R. Mcintyre, Y. Lee, The efficacy of mind-body (Baduanjin) exercise on self-reported sleep quality and quality of life in elderly subjects with sleep disturbances: a randomized controlled trial, Sleep Breath, 24(2) (2020) 695–701.

[112] W.Y. Yue, J.M. Cao, H.T. Zhou, R.M. Xu, Tai Chi in combination with acupoint massage can improve sleep quality of elderly patients with chronic insomnia, Int. J. Clin. Exp. Med., 9(2) (2016) 4316–4323.

[113] F.C. Lai, I.H. Chen, P.J. Chen, I.J. Chen, H.W. Chien, C.F. Yuan, Acupressure, Sleep, and Quality of Life in Institutionalized Older Adults: A Randomized Controlled Trial, JOURNAL OF THE AMERICAN GERIATRICS SOCIETY, 65(5) (2017) E103–E108.

[114] H.Y. Chiu, H.C. Lee, J.W. Liu, S.J. Hua, P.Y. Chen, P.S. Tsai, Y.K. Tu, Comparative efficacy and safety of hypnotics for insomnia in older adults: a systematic review and network meta-analysis, Sleep, 44(5) (2021).

[115] C.Y. Kwon, B. Lee, M.J. Cheong, T.H. Kim, B.H. Jang, S.Y. Chung, J.W. Kim, Non-pharmacological Treatment for Elderly Individuals With Insomnia: A Systematic Review and Network Meta-Analysis, Front Psychiatry, 11 (2020) 608896.

[116] M.T. Samara, M. Huhn, V. Chiocchia, J. Schneider-Thoma, M. Wiegand, G. Salanti, S. Leucht, Efficacy, acceptability, and tolerability of all available treatments for insomnia in the elderly: a systematic review and network meta-analysis, Acta Psychiatr Scand, 142(1) (2020) 6–17.

[117] F. Feng, S. Tuchman, J.W. Denninger, G.L. Fricchione, A. Yeung, Qigong for the Prevention, Treatment, and Rehabilitation of COVID-19 Infection in Older Adults, Am J Geriatr Psychiatry, 28(8) (2020) 812-819.

[118] D. Qi, N.M.L. Wong, R. Shao, I.S.C. Man, C.H.Y. Wong, L.P. Yuen, C.C.H. Chan, T.M.C. Lee, Qigong exercise enhances cognitive functions in the elderly via an interleukin-6-hippocampus pathway: A randomized active-controlled trial, Brain Behav Immun, 95 (2021) 381–390.

[119] P.M. Siu, A.P. Yu, B.T. Tam, E.C. Chin, D.S. Yu, K.F. Chung, S.S. Hui, J. Woo, D.Y. Fong, P.H. Lee, G.X. Wei, M.R. Irwin, Effects of Tai Chi or Exercise on Sleep in Older Adults With Insomnia: A Randomized Clinical Trial, JAMA Netw Open, 4(2) (2021) e2037199.

[120] P.S. Chang, T. Knobf, B. Oh, M. Funk, Physical and Psychological Health Outcomes of Qigong Exercise in Older Adults: A Systematic Review and Meta-Analysis, Am J Chin Med, 47(2) (2019) 301–322.

[121] M. Johansson, P. Hassmén, Acute psychological responses to Qigong exercise of varying durations, Am J Chin Med, 36(3) (2008) 449–58.

[122] C.Y. Lin, T.T. Wei, C.C. Wang, W.C. Chen, Y.M. Wang, S.Y. Tsai, Acute Physiological and Psychological Effects of Qigong Exercise in Older Practitioners, Evid Based Complement Alternat Med, 2018 (2018) 4960978.

[123] M.S. Chin, S.N. Kales, Understanding mind-body disciplines: A pilot study of paced breathing and dynamic muscle contraction on autonomic nervous system reactivity, Stress Health, 35(4) (2019) 542–548.

[124] R. Jahnke, L. Larkey, C. Rogers, J. Etnier, F. Lin, A comprehensive review of health benefits of qigong and tai chi, Am J Health Promot, 24(6) (2010) e1–e25.

