## Supplementary for "Comparative efficacy of Qigong exercise and other external treatments of traditional Chinese medicine on insomnia in the elderly: A network meta-analysis"

Content

### Appendix 1 Search Strategy and Results

**Number of citations by each database and trial register searched***

| **Databases and Trial registers** | **Citations** |
| --- | --- |
| **Databases：** |  |
| Pubmed | 1089 |
| Web of Science | 1807 |
| Embase | 1302 |
| CNKI | 4710 |
| WanFang Data | 2978 |
| VIP | 1914 |
| **Total (databases)** | **13800** |

**Full search strategy for each database**

**PubMed**

#1 Search: "Sleep Initiation and Maintenance Disorders"[Mesh]

#2 Search:(((((((((((((((((((((((Primary insomnia[Title/Abstract]) OR (Nonorganic Insomnia[Title/Abstract])) OR (Chronic Insomnia[Title/Abstract])) OR (Sleepless[Title/Abstract])) OR (Sleeplessness[Title/Abstract])) OR (Sleep Initiation and Maintenance Disorders [Title/Abstract])) OR (Insomnia*[Title/Abstract])) OR (Transient Insomnia[Title/Abstract])) OR (Insomnia Disorder*[Title/Abstract])) OR (DIMS[Title/Abstract])) OR (Insomnia, Transient[Title/Abstract])) OR (Insomnia, Chronic[Title/Abstract])) OR (Disorders of Initiating and Maintaining Sleep [Title/Abstract])) OR (Rebound Insomnia[Title/Abstract])) OR (Psychophysiological Insomnia[Title/Abstract])) OR (Early Awakening[Title/Abstract])) OR (Insomnia, Rebound[Title/Abstract])) OR (Insomnia, Psychophysiological[Title/Abstract])) OR (Awakening, Early[Title/Abstract])) OR (Secondary Insomnia[Title/Abstract])) OR (Sleep Disorders[Title/Abstract])) OR (Insomnia, Primary[Title/Abstract])) OR (Insomnia, Secondary[Title/Abstract])) OR (Insomnia, Nonorganic[Title/Abstract]) OR (Sleep Initiation Dysfunction*[Title/Abstract]) OR (Dysfunction*, Sleep Initiation[Title/Abstract])

#3 #1 OR #3

#4	Search: (((((((((((("Medicine, Chinese Traditional"[Mesh]) OR ("Complementary Therapies"[Mesh])) OR ("Iontophoresis"[Mesh])) OR ("Auriculotherapy"[Mesh])) OR ("Acupuncture, Ear"[Mesh])) OR ("Acupuncture"[Mesh] OR "Acupuncture Therapy"[Mesh])) OR (("Electroacupuncture"[Mesh]) OR "Acupressure"[Mesh])) OR ("Moxibustion"[Mesh])) OR ("Cupping Therapy"[Mesh])) OR ("Massage"[Mesh])) OR ("Qigong"[Mesh])) OR ("Tai Ji"[Mesh])) OR ("Breathing Exercises"[Mesh])

#5 Search:((((((((((((Traditional Chinese Medicine[Title/Abstract]) OR (Chinese Traditional Medicine[Title/Abstract])) OR (Chinese Medicine, Traditional[Title/Abstract])) OR (Chung I Hsueh[Title/Abstract])) OR (Traditional Medicine, Chinese[Title/Abstract])) OR (Medicine,Chinese Traditional[Title/Abstract])) OR (Hsueh, Chung I[Title/Abstract])) OR (Zhong Yi Xue[Title/Abstract])) OR (Complementary Medicine[Title/Abstract])) OR (Alternative Medicine[Title/Abstract])) OR (alternative therap*[Title/Abstract])) OR (Complementary Therap*[Title/Abstract])) OR (Iontophore*[Title/Abstract])

#6 Search: (((((((((((((((((((Ear + Acupunctur*[Title/Abstract]) OR (Auricular + Acupunctur*[Title/Abstract])) OR (Auricular + Acupressur*[Title/Abstract])) OR (Auriculotherap*[Title/Abstract])) OR (Auricular + Poin*[Title/Abstract])) OR (Auricular + Plaste*[Title/Abstract])) OR (Ear + Poin*[Title/Abstract])) OR (Ear + Acupoin*[Title/Abstract])) OR (Ear + Acupressur*[Title/Abstract])) OR (Otopoin*[Title/Abstract])) OR (Auricular + Massag*[Title/Abstract])) OR (Ear + Massag*[Title/Abstract])) OR (Ear Hol*[Title/Abstract])) OR (Vaccaria*[Title/Abstract])) OR (Seed*[Title/Abstract])) OR (Magne*+ Auricu*[Title/Abstract])) OR (Magne*+ Ear[Title/Abstract])) OR (Erxue[Title/Abstract])) OR (Auriculotherapy[Title/Abstract])) OR (Acupuncture,Ear[Title/Abstract])

#7 Search:(((((((((((((((((((((((((Acupuncture[Title/Abstract]) OR (Acupuncture Therapy[Title/Abstract])) OR (Electroacupuncture[Title/Abstract])) OR (Acupressure[Title/Abstract])) OR (Acupuncture Treatment*[Title/Abstract])) OR (Therapy, Pharmacoacupuncture[Title/Abstract])) OR (Treatment, Acupuncture[Title/Abstract])) OR (Therapy, Acupuncture[Title/Abstract])) OR (Pharmacoacupuncture Treatment[Title/Abstract])) OR (Treatment, Pharmacoacupuncture[Title/Abstract])) OR (Pharmacoacupuncture Therapy[Title/Abstract])) OR (Acupotom*[Title/Abstract])) OR (Pharmacopuncture[Title/Abstract])) OR (electroacupunct*[Title/Abstract])) OR (acupuncture[Title/Abstract] OR moxibustion[Title/Abstract])) OR (Shiatsu[Title/Abstract])) OR (Zhi Ya[Title/Abstract])) OR (Chih Ya[Title/Abstract])) OR (Auricular Acupunture[Title/Abstract])) OR (acupunct*[Title/Abstract])) OR (acupress*[Title/Abstract])) OR (acupoint*[Title/Abstract])) OR (moxibust*[Title/Abstract])) OR (electro‐acupunct*[Title/Abstract])) OR (auriculotherap*[Title/Abstract])) OR (auriculoacupunct*[Title/Abstract])

#8 Search:((((((Moxibustion[Title/Abstract]) OR (Moxabustion therapy[Title/Abstract])) OR (Moxabustion[Title/Abstract])) OR (warm* acupuncture[Title/Abstract])) OR (Needl* Warm*[Title/Abstract])) OR (Moxabustion Treatment[Title/Abstract])) OR (Moxa[Title/Abstract])

#9 Search: ((((((((((((((((((acupoint injection[Title/Abstract]) OR (acupoint application[Title/Abstract])) OR (scraping therapy[Title/Abstract])) OR (Guasha treatment[Title/Abstract])) OR (Scraping sha treatment[Title/Abstract])) OR (CUPPING THERAPY[Title/Abstract])) OR (Cupping Therapies[Title/Abstract])) OR (Therapy, Cupping[Title/Abstract])) OR (Cupping Treatment*[Title/Abstract])) OR (Treatment, Cupping[Title/Abstract])) OR (Cupping[Title/Abstract])) OR (Massage[Title/Abstract])) OR (Zone Therap*[Title/Abstract])) OR (Therap*, Zone[Title/Abstract])) OR (Massage Therap*[Title/Abstract])) OR (Therap*, Massage[Title/Abstract])) OR (Tuina[Title/Abstract])) OR (Qigong[Title/Abstract])) OR (Ch'i Kung[Title/Abstract])

#10 Search:(((((((((((((((((((((((((Tai Chi[Title/Abstract]) OR (Chi, Tai[Title/Abstract])) OR (Tai Ji Quan[Title/Abstract])) OR (Ji Quan, Tai[Title/Abstract])) OR (Quan, Tai Ji[Title/Abstract])) OR (Taiji[Title/Abstract])) OR (Taijiquan[Title/Abstract])) OR (T'ai Chi[Title/Abstract])) OR (Tai Chi Chuan[Title/Abstract])) OR (SHADOW BOXING[Title/Abstract])) OR (Tai Ji[Title/Abstract])) OR (Tai-ji[Title/Abstract])) OR (wuqinxi[Title/Abstract])) OR (Baduanjin[Title/Abstract])) OR (eight section brocades[Title/Abstract])) OR (Liuzijue[Title/Abstract])) OR (Mawangdui Daoyin[Title/Abstract])) OR (Daoyin*[Title/Abstract])) OR (Ch'i Kung[Title/Abstract])) OR (Health Qigong[Title/Abstract])) OR (qigong[Title/Abstract])) OR (Exercise, Breathing[Title/Abstract])) OR (Respiratory Muscle Training[Title/Abstract])) OR (Muscle Training, Respiratory[Title/Abstract])) OR (Training, Respiratory Muscle[Title/Abstract])) OR (BREATHING EXERCISES[Title/Abstract])

#11 Search:(((((five-element music[Title/Abstract]) OR (five lines of music of Chinese medicine[Title/Abstract])) OR (five-tone therapy[Title/Abstract])) OR (acupuncture point paste[Title/Abstract])) OR (Chinese herbal soaking[Title/Abstract])) OR (external treatments of traditional Chinese medicine[Title/Abstract])

#12 #4 OR #5 OR #6 OR #7 OR #8 OR #9 OR #10 OR#11

#13 Search: ((("Randomized Controlled Trial" [Publication Type]) OR "Controlled Clinical Trial" [Publication Type])) OR (((((((randomized[Title/Abstract]) OR randomised[Title/Abstract]) OR placebo[Title/Abstract]) OR sham[Title/Abstract]) OR randomly[Title/Abstract]) OR trial[Title/Abstract]) OR groups[Title/Abstract])

#14 Search: ((animals[MeSH Terms] NOT (humans[MeSH Terms] AND animals[MeSH Terms])))

#15 #13 NOT#14

#16 #3 AND #12 AND #15

**Web of science**

#1 TS=(((((((((((((((((((((((((Primary insomnia) OR (Nonorganic Insomnia)) OR (Chronic Insomnia)) OR (Sleepless)) OR (Sleeplessness)) OR (Sleep Initiation and Maintenance Disorders)) OR (Insomnia*)) OR (Transient Insomnia)) OR (Insomnia Disorder*)) OR (DIMS)) OR (Insomnia, Transient)) OR (Insomnia, Chronic)) OR (Disorders of Initiating and Maintaining Sleep)) OR (Rebound Insomnia)) OR (Psychophysiological Insomnia)) OR (Early Awakening)) OR (Insomnia, Rebound)) OR (Insomnia, Psychophysiological)) OR (Awakening, Early)) OR (Secondary Insomnia)) OR (Sleep Disorders)) OR (Insomnia, Primary)) OR (Insomnia, Secondary)) OR (Insomnia, Nonorganic)) OR (Sleep Initiation Dysfunction*)) OR (Dysfunction*, Sleep Initiation)

#2 TS=(((((((((((((((((((("Medicine, Chinese Traditional") OR ("Complementary Therapies")) OR ("Iontophoresis")) OR ("Auriculotherapy")) OR ("Acupuncture, Ear")) OR ("Acupuncture" ))OR ("Acupuncture Therapy")) OR (("Electroacupuncture") OR "Acupressure")) OR ("Moxibustion")) OR ("Cupping Therapy")) OR ("Massage")) OR ("Qigong")) OR ("Tai Ji")) OR ("Breathing Exercises")) OR (((((((((((((Traditional Chinese Medicine) OR (Chinese Traditional Medicine)) OR (Chinese Medicine, Traditional)) OR (Chung I Hsueh)) OR (Traditional Medicine, Chinese)) OR (Medicine,Chinese Traditional)) OR (Hsueh, Chung I)) OR (Zhong Yi Xue)) OR (Complementary Medicine)) OR (Alternative Medicine)) OR (alternative therap*)) OR (Complementary Therap*)) OR (Iontophore*))) OR ((((((((((((((((((((Ear + Acupunctur*) OR (Auricular + Acupunctur*)) OR (Auricular + Acupressur*)) OR (Auriculotherap*)) OR (Auricular + Poin*)) OR (Auricular + Plaste*)) OR (Ear + Poin*)) OR (Ear + Acupoin*)) OR (Ear + Acupressur*)) OR (Otopoin*)) OR (Auricular + Massag*)) OR (Ear + Massag*)) OR (Ear Hol*)) OR (Vaccaria*)) OR (Seed*)) OR (Magne*+ Auricu*)) OR (Magne*+ Ear)) OR (Erxue)) OR (Auriculotherapy)) OR (Acupuncture,Ear))) OR ((((((((((((((((((((((((((Acupuncture) OR (Acupuncture Therapy)) OR (Electroacupuncture)) OR (Acupressure)) OR (Acupuncture Treatment*)) OR (Therapy, Pharmacoacupuncture)) OR (Treatment, Acupuncture)) OR (Therapy, Acupuncture)) OR (Pharmacoacupuncture Treatment)) OR (Treatment, Pharmacoacupuncture)) OR (Pharmacoacupuncture Therapy)) OR (Acupotom*)) OR (Pharmacopuncture)) OR (electroacupunct*)) OR (acupuncture OR moxibustion)) OR (Shiatsu)) OR (Zhi Ya)) OR (Chih Ya)) OR (Auricular Acupunture)) OR (acupunct*)) OR (acupress*)) OR (acupoint*)) OR (moxibust*)) OR (electro‐acupunct*)) OR (auriculotherap*)) OR (auriculoacupunct*))) OR ((((((((Moxibustion) OR (Moxa)) OR (Moxabustion therapy)) OR (Moxabustion)) OR (warm* acupuncture)) OR (Needl* Warm*)) OR (Moxabustion Treatment)) OR (Moxa))) OR (((((((((((((((((((acupoint injection) OR (acupoint application)) OR (scraping therapy)) OR (Guasha treatment)) OR (Scraping sha treatment)) OR (CUPPING THERAPY)) OR (Cupping Therapies)) OR (Therapy, Cupping)) OR (Cupping Treatment*)) OR (Treatment, Cupping)) OR (Cupping)) OR (Massage)) OR (Zone Therap*)) OR (Therap*, Zone)) OR (Massage Therap*)) OR (Therap*, Massage)) OR (Tuina)) OR (Qigong)) OR (Ch'i Kung))) OR ((((((((((((((((((((((((((((Tai Chi) OR (Tai Ji)) OR (Chi, Tai)) OR (Tai Ji Quan)) OR (Ji Quan, Tai)) OR (Quan, Tai Ji)) OR (Taiji)) OR (Taijiquan)) OR (T'ai Chi)) OR (Tai Chi Chuan)) OR (SHADOW BOXING)) OR (Tai Ji)) OR (Tai-ji)) OR (wuqinxi)) OR (Baduanjin)) OR (eight section brocades)) OR (Liuzijue)) OR (BREATHING EXERCISES)) OR (Mawangdui Daoyin)) OR (Daoyin*)) OR (Ch'i Kung)) OR (Health Qigong)) OR (qigong)) OR (Exercise, Breathing)) OR (Respiratory Muscle Training)) OR (Muscle Training, Respiratory)) OR (Training, Respiratory Muscle)) OR (BREATHING EXERCISES))) OR ((((((five-element music) OR (five lines of music of Chinese medicine)) OR (five-tone therapy)) OR (acupuncture point paste)) OR (Chinese herbal soaking)) OR (external treatments of traditional Chinese medicine))

#3 TS=(random* OR crossover* OR (cross NEAR/3 over*) OR placebo OR (doubl* NEAR/3 blind*) OR (doubl* NEAR/3 mask*) OR (singl* NEAR/3 blind*) OR (singl* NEAR/3 mask*) OR (trebl* NEAR/3 blind*) OR (trebl* NEAR/3 mask*) OR (tripl* NEAR/3 blind*) OR (tripl* NEAR/3 mask*) OR assign* OR allocat* OR volunteer*)

#4 #1 AND #2 AND #3

**EMBASE**

#1 'primary insomnia'/exp

#2 'chronic insomnia'/exp

#3 'insomnia'/exp

#4 'primary insomnia':ab,ti OR 'nonorganic insomnia':ab,ti OR 'chronic insomnia':ab,ti OR sleepless:ab,ti OR sleeplessness:ab,ti OR 'sleep initiation':ab,ti OR 'maintenance disorders':ab,ti OR insomnia*:ab,ti OR 'transient insomnia':ab,ti OR 'insomnia disorder*':ab,ti OR dims:ab,ti OR 'disorders of initiating':ab,ti OR 'maintaining sleep':ab,ti OR 'insomnia, transient':ab,ti OR 'insomnia, chronic':ab,ti OR 'rebound insomnia':ab,ti OR 'psychophysiological insomnia':ab,ti OR 'early awakening':ab,ti OR 'insomnia, rebound':ab,ti OR 'insomnia, psychophysiological':ab,ti OR 'awakening, early':ab,ti OR 'secondary insomnia':ab,ti OR 'sleep disorders':ab,ti OR 'insomnia, primary':ab,ti OR 'insomnia, secondary':ab,ti OR 'insomnia, nonorganic':ab,ti OR 'sleep initiation dysfunction*':ab,ti OR 'dysfunction*, sleep initiation':ab,ti

#5	#1 OR #2 OR #3 OR #4

#6	'chinese medicine'/exp

#7	'alternative medicine'/exp

#8	'iontophoresis'/exp

#9	'auricular acupuncture'/exp

#10 'acupuncture'/exp

#11 'electroacupuncture'/exp

#12 'acupressure'/exp

#13 'moxibustion'/exp

#14 'cupping therapy'/exp

#15 'massage'/exp

#16 'qigong'/exp

#17 'tai chi'/exp

#18 'breathing exercise'/exp

#19 'medicine, chinese traditional':ab,ti OR 'complementary therapies':ab,ti OR 'iontophoresis':ab,ti OR 'auriculotherapy':ab,ti OR 'acupuncture, ear':ab,ti OR 'acupuncture':ab,ti OR 'electroacupuncture':ab,ti OR 'acupressure':ab,ti OR 'moxibustion':ab,ti OR 'massage':ab,ti OR 'qigong':ab,ti OR 'traditional chinese medicine':ab,ti OR 'chinese traditional medicine':ab,ti OR 'chinese medicine, traditional':ab,ti OR 'chung i hsueh':ab,ti OR 'traditional medicine, chinese':ab,ti OR 'medicine,chinese traditional':ab,ti OR 'hsueh, chung i':ab,ti OR 'zhong yi xue':ab,ti OR 'complementary medicine':ab,ti OR 'alternative medicine':ab,ti OR 'alternative therap*':ab,ti OR 'complementary therap*':ab,ti OR iontophore*:ab,ti OR 'ear + acupunctur*':ab,ti OR 'auricular + acupunctur*':ab,ti OR 'auricular + acupressur*':ab,ti OR 'auricular + poin*':ab,ti OR 'auricular + plaste*':ab,ti OR 'ear + poin*':ab,ti OR 'ear + acupoin*':ab,ti OR 'ear + acupressur*':ab,ti OR otopoin*:ab,ti OR 'auricular + massag*':ab,ti OR 'ear + massag*':ab,ti OR 'ear hol*':ab,ti OR vaccaria*:ab,ti OR seed*:ab,ti OR 'magne*+ auricu*':ab,ti OR 'magne*+ ear':ab,ti OR erxue:ab,ti OR auriculotherapy:ab,ti OR acupuncture,ear:ab,ti OR 'acupuncture therapy':ab,ti OR electroacupuncture:ab,ti OR acupressure:ab,ti OR 'acupuncture treatment*':ab,ti OR 'therapy, pharmacoacupuncture':ab,ti OR 'treatment, acupuncture':ab,ti OR 'therapy, acupuncture':ab,ti OR 'pharmacoacupuncture treatment':ab,ti OR 'treatment, pharmacoacupuncture':ab,ti OR 'pharmacoacupuncture therapy':ab,ti OR acupotom*:ab,ti OR pharmacopuncture:ab,ti OR electroacupunct*:ab,ti OR acupuncture:ab,ti OR shiatsu:ab,ti OR 'zhi ya':ab,ti OR 'chih ya':ab,ti OR 'auricular acupunture':ab,ti OR acupunct*:ab,ti OR acupress*:ab,ti OR acupoint*:ab,ti OR moxibust*:ab,ti OR electro‐acupunct*:ab,ti OR auriculotherap*:ab,ti OR auriculoacupunct*:ab,ti OR moxibustion:ab,ti OR 'moxabustion therapy':ab,ti OR moxabustion:ab,ti OR 'warm* acupuncture':ab,ti OR 'needl* warm*':ab,ti OR 'moxabustion treatment':ab,ti OR moxa:ab,ti OR 'acupoint injection':ab,ti OR 'acupoint application':ab,ti OR 'scraping therapy':ab,ti OR 'guasha treatment':ab,ti OR 'scraping sha treatment':ab,ti OR 'cupping therapy':ab,ti OR 'cupping therapies':ab,ti OR 'therapy, cupping':ab,ti OR 'cupping treatment*':ab,ti OR 'treatment, cupping':ab,ti OR cupping:ab,ti OR massage:ab,ti OR 'zone therap*':ab,ti OR 'therap*, zone':ab,ti OR 'massage therap*':ab,ti OR 'therap*, massage':ab,ti OR tuina:ab,ti OR 'chi, tai':ab,ti OR 'tai ji quan':ab,ti OR 'ji quan, tai':ab,ti OR 'quan, tai ji':ab,ti OR taiji:ab,ti OR taijiquan:ab,ti OR 'tai chi':ab,ti OR 'tai chi chuan':ab,ti OR 'shadow boxing':ab,ti OR 'tai ji':ab,ti OR wuqinxi:ab,ti OR baduanjin:ab,ti OR 'eight section brocades':ab,ti OR liuzijue:ab,ti OR 'mawangdui daoyin':ab,ti OR daoyin*:ab,ti OR 'chi kung':ab,ti OR 'health qigong':ab,ti OR qigong:ab,ti OR 'exercise, breathing':ab,ti OR 'respiratory muscle training':ab,ti OR 'muscle training, respiratory':ab,ti OR 'training, respiratory muscle':ab,ti OR 'breathing exercises':ab,ti OR 'five-element music':ab,ti OR 'five lines of music of chinese medicine':ab,ti OR 'five-tone therapy':ab,ti OR 'acupuncture point paste':ab,ti OR 'chinese herbal soaking':ab,ti OR 'external treatments of traditional chinese medicine':ab,ti

#20 #6 OR #7 OR #8 OR #9 OR #10 OR #11 OR #12 OR #13 OR #14 OR #15 OR #16 OR #17 OR #18

#21 #19 OR #20

#22 #5 AND #21

#23 random*:ab,ti OR crossover*:ab,ti OR ((cross NEAR/3 over*):ab,ti) OR placebo:ab,ti OR ((doubl* NEAR/3 blind*):ab,ti) OR ((doubl* NEAR/3 mask*):ab,ti) OR ((singl* NEAR/3 blind*):ab,ti) OR ((singl* NEAR/3 mask*):ab,ti) OR ((trebl* NEAR/3 blind*):ab,ti) OR ((trebl* NEAR/3 mask*):ab,ti) OR ((tripl* NEAR/3 blind*):ab,ti) OR ((tripl* NEAR/3 mask*):ab,ti) OR assign*:ab,ti OR allocat*:ab,ti OR volunteer*:ab,ti

#24 'controlled clinical trial'/exp OR 'single blind procedure'/exp OR 'double-blind procedure'/exp OR 'crossover procedure'/exp

#25 #23 OR #24

#26 'animal'/exp OR 'nonhuman'/exp OR 'animal experiment'/exp

#27 'human'/exp

#28 #26 AND #27

#29 #26 NOT #28

#30 #25 NOT #29

#31 #22 AND #

### Appendix 2 The risk of bias for each study

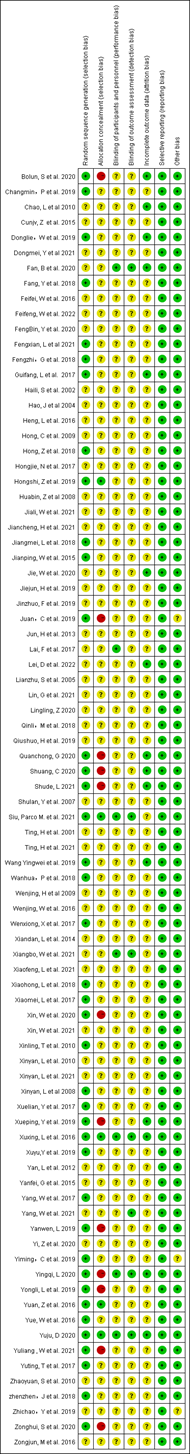

Figure 1 The risk of bias for each study

### Appendix 3 Results from pairwise meta-analysis for each outcome

#### Appendix 3.1 Pairwise analysis for the total effective (TE) rate

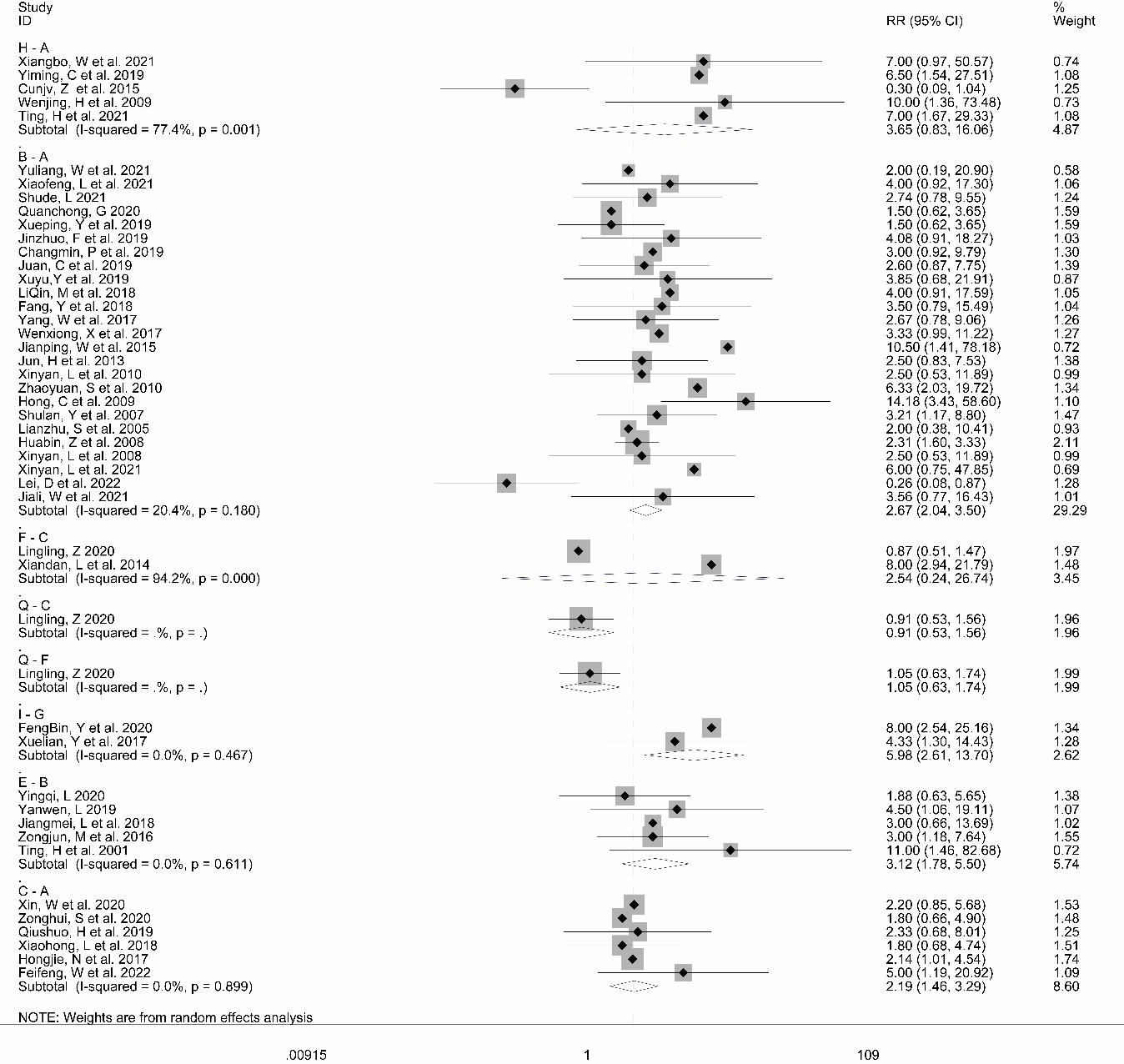

Figure 2 Pairwise analysis for the total effective (TE) rate

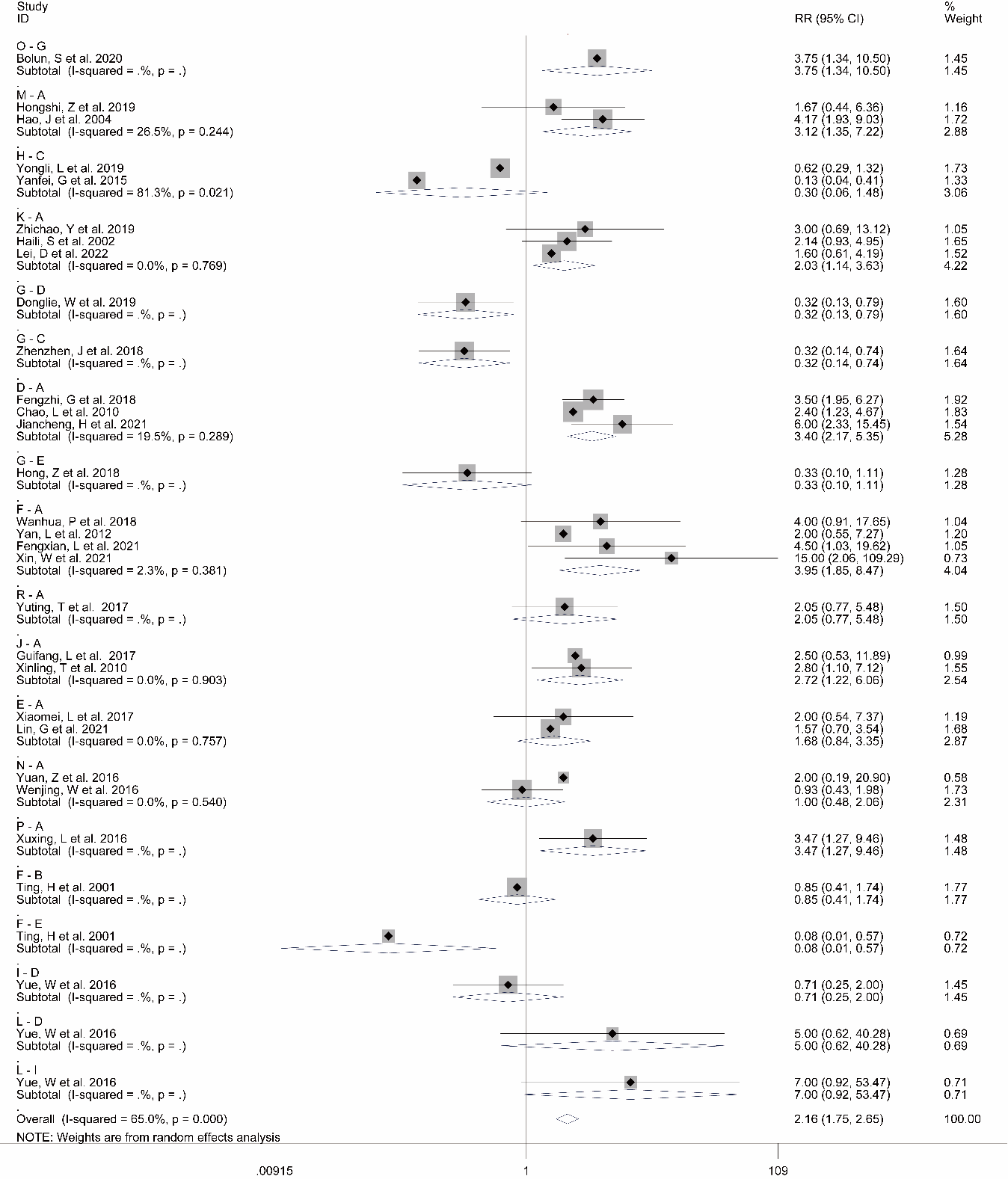

(Figure 2 continues on next page)

Note. A=RT; B=ACP; C=CT; D=Qigong; E=AAM; F=AT; G=HE; H=HMFB; I=AM; J=Moxibustion; K=APAM; L=QAM; M=MT; N=CHF; O=MAM; P=FEM; Q=AST; R=CMIT

#### Appendix 3.2 Pairwise analysis for Pittsburgh Sleep Quality Index (PSQI) scores

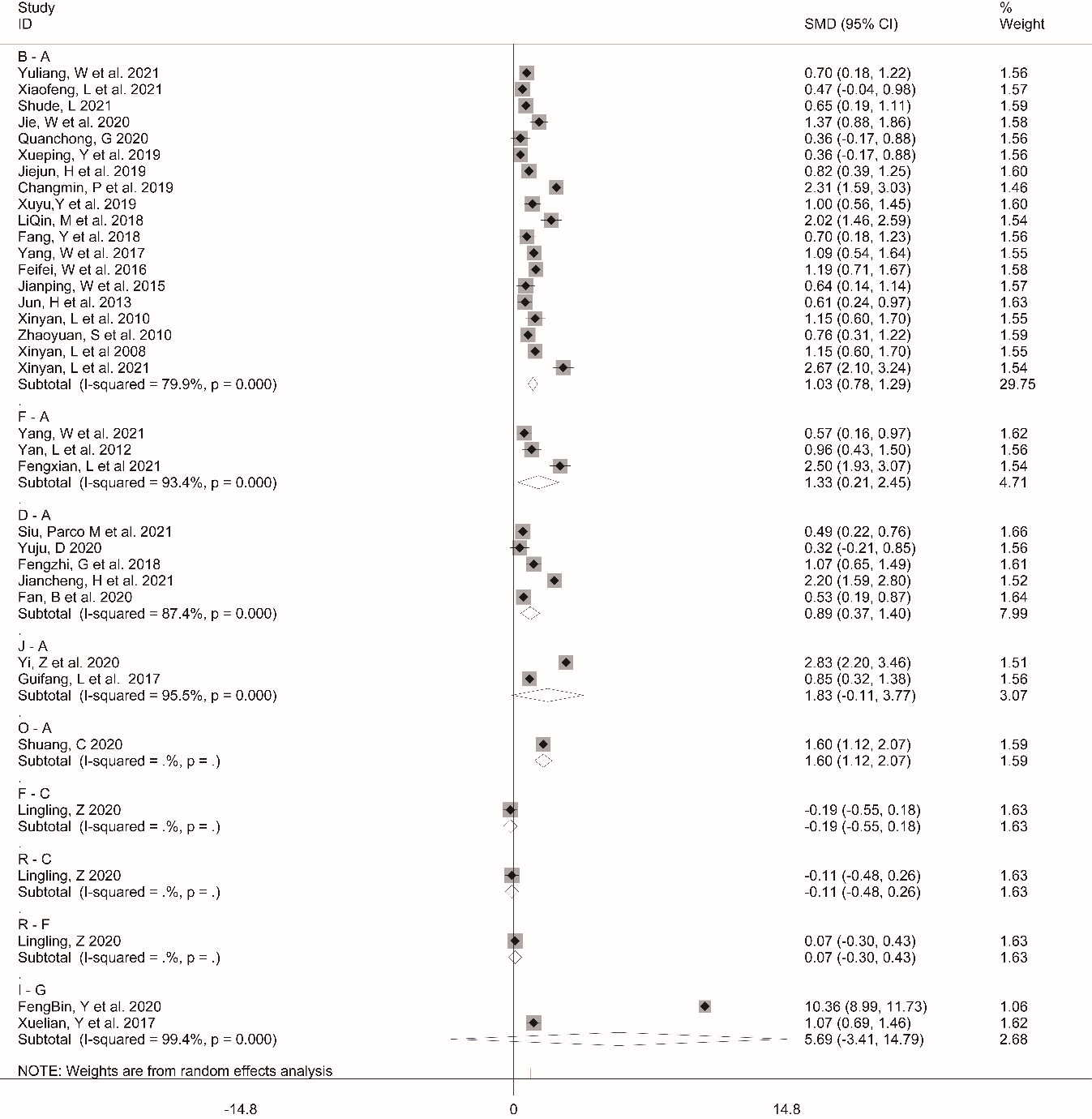

Figure 3 Pairwise analysis for Pittsburgh Sleep Quality Index (PSQI) scores

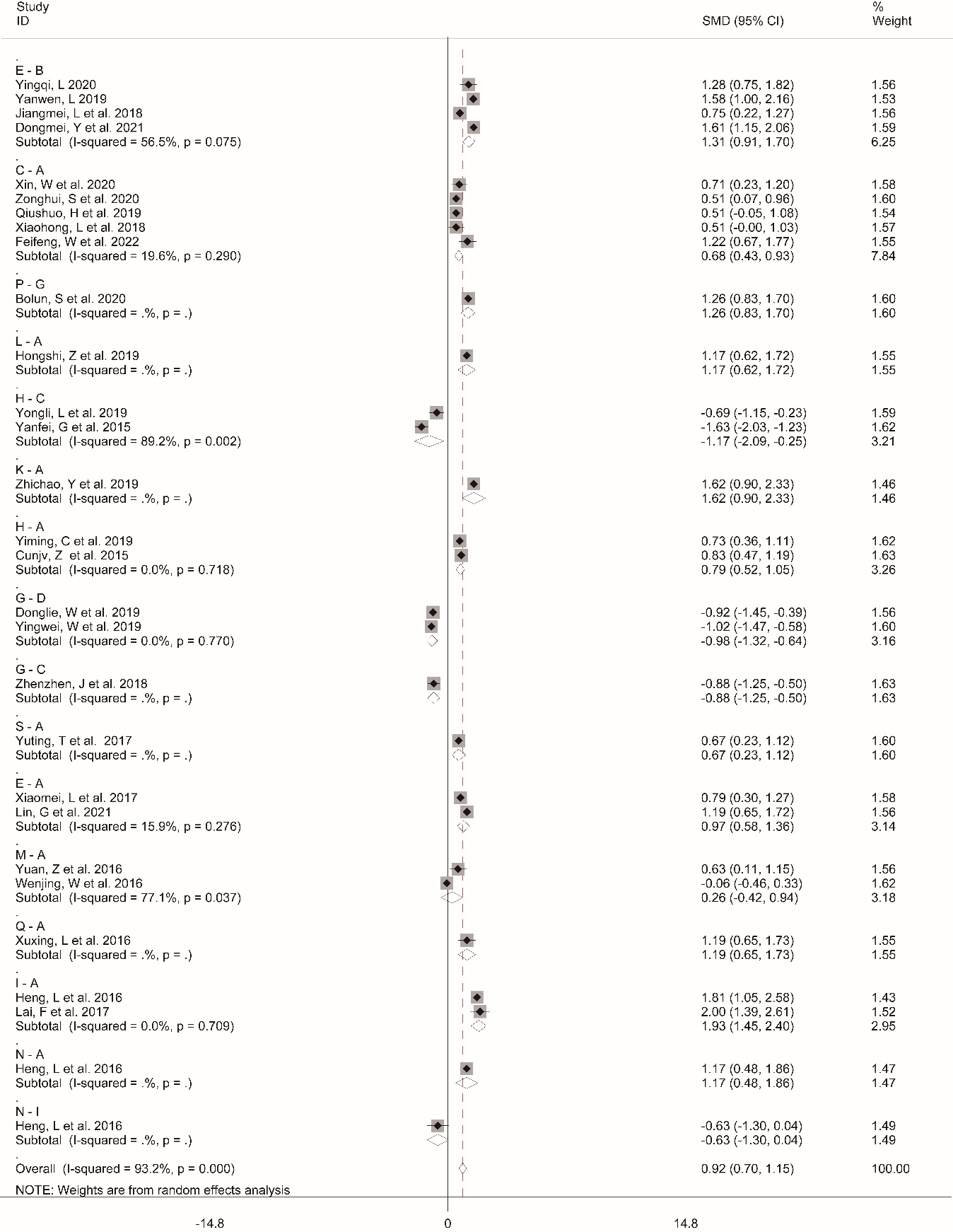

(Figure 3 continues on next page)

Note. A=RT; B=ACP; C=CT; D=Qigong; E=AAM; F=AT; G=HE; H=HMFB; I=AM; J=Moxibustion; K=APAM; L=MT; M=CHF; N=Aromatherapy; O=Scrapie; P=MAM; Q=FEM; R=AST; S=CMIT

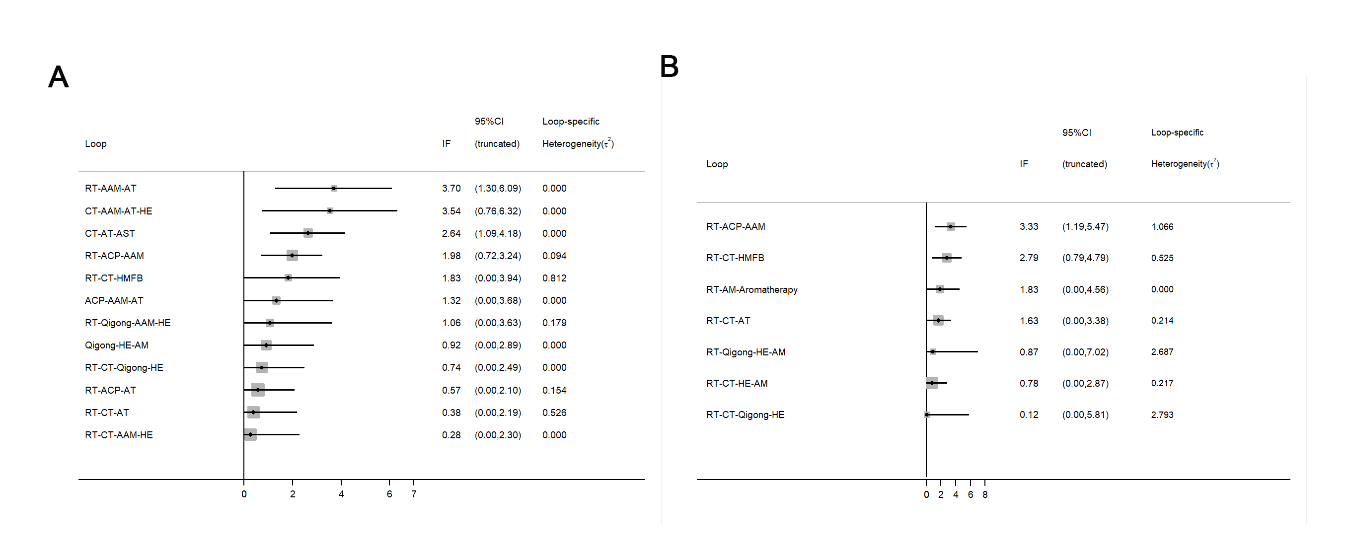

### Appendix 4 Loop-specific heterogeneity for each outcome

Figure 4 Loop-specific heterogeneity for each outcome

*Note.* A: the total effective (TE) rate; B: Pittsburgh Sleep Quality Index (PSQI) scores

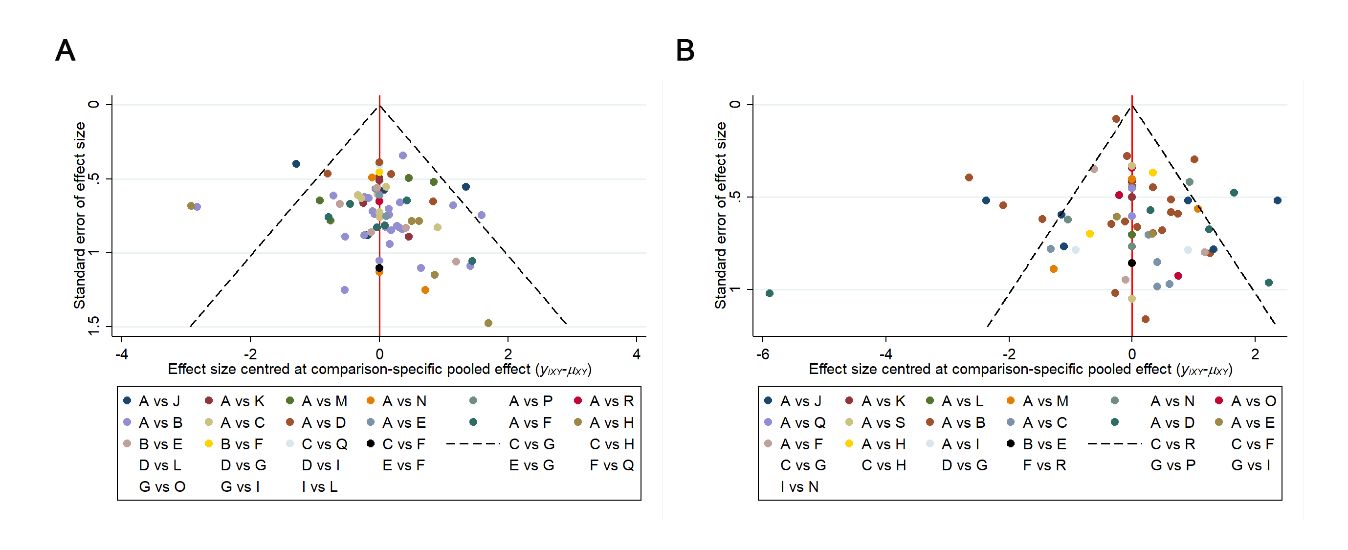

### Appendix 5 Funnel plot for each outcome from the network meta-analysis

Figure 5

*Note.* A: the total effective (TE) rate; B: Pittsburgh Sleep Quality Index (PSQI) scores
