## Supplementary material for "Comparative efficacy of Qigong exercise and other external treatments of traditional Chinese medicine on insomnia in the elderly: A network meta-analysis": Tables

**Table 1** Characteristics of included studies

| **Author** | **Year Published** | **Study setting** | **Study design** | **Participants'age (mean(SD) or ranges)(years)** | **N (%) Men** | **Participants (n)** | **Interventions** | **Comparator** | **Intervention duration (months)** | **Intervention frequency** | **Outcomes assessed** |
| --- | --- | --- | --- | --- | --- | --- | --- | --- | --- | --- | --- |
| Xiangbo, W et al. [29] | 2021 | Guangdong, China | Randomized, patient-, analyst-blinded, controlled study | INT=71.58±4.16; CON=71.61±4.12 | INT=9 (56.3); CON=8 (50.0) | Randomized=32; Completed=32; INT=16; CON=16 | HMFB | RT | 1 | Once a day | TE rate; PSQI |
| Yuliang, W et al. [30] | 2021 | Xinjiang, China | Prospective randomized, controlled study | INT=63.39±4.732; CON=64.38±5.342 | INT=14 (46.7); CON=12 (40.0) | Randomized=60; Completed=60; INT=30; CON=30 | ACP | RT | 0.7 | Once a day, after six days of treatment, one day off | TE rate; PSQI |
| Yang, W et al. [31] | 2021 | Jilin, China | Prospective randomized, controlled study | INT=66.19±10.67; CON=65.32±11.31 | INT=29 (59.2); CON=28 (57.1) | Randomized=98; Completed=98; INT=49; CON=49 | AT | RT | 2 | Once a day | PSQI |
| Xiaofeng, L et al. [32] | 2021 | Guangxi, China | Prospective randomized, controlled study | INT=72.98±3.21; CON=71.99±2.09 | INT=16 (53.3); CON=14 (46.7) | Randomized=60; Completed=60; INT=30; CON=30 | ACP | RT | 0.33 | Once a day | TE rate; PSQI |
| Shude, L[33] | 2021 | Fujian, China | Prospective randomized, controlled study | INT=66.29±4.96; CON=65.62±3.72 | INT=17 (43.6); CON=20 (52.6) | Randomized=80; Completed=77; INT=39; CON=38 | ACP | RT | 1 | Once a day | TE rate; PSQI |
| Siu, Parco M et al. [34] | 2021 | HongKong, China | Randomized, 3-arm, parallel, assessor-masked clinical trail | INT=66.5(6.4); CON=68.0(8.2) | INT=21(20.0); CON=22(20.0) | Randomized=320; Completed=320; INT=105; CON=110 | Qigong | RT | 3 | Three times a week | PSQI |
| Jie, W et al. [35] | 2020 | Shandong, China | Prospective randomized, controlled study | INT=64.03±2.87; CON=64.93±3.28 | INT=46 (57.5); CON=39 (48.7) | Randomized=160; Completed=80; INT=40; CON=40 | ACP | RT | 1.3 | Once a day | PSQI |
| Quanchong, G[36] | 2020 | Heilongjiang, China | Prospective randomized, controlled study | INT=71.3±5.7; CON=72.3±4.8 | INT=15 (53.6); CON=12 (42.9) | Randomized=60; Completed=56; INT=28; CON=28 | ACP | RT | 1 | Once a day | TE rate; PSQI |
| Yi, Z et al. [37] | 2020 | Guangxi, China | Prospective randomized, controlled study | INT=73.2±3.3; CON=72.2±2.3 | INT=18 (45); CON=17 (44.7) | Randomized=78; Completed=78; INT=40; CON=38 | Moxibustion | RT | 0.67 | Once a day | PSQI |
| Shuang, C[38] | 2020 | Guangzhou, China | Prospective randomized, controlled study | INT=67.00±4.83; CON=66.63±3.95 | INT=14 (46.7); CON=12 (40.1) | Randomized=92; Completed=91; INT=45; CON=46 | Scrapie | RT | 1.5 | Once a week | PSQI |
| Yuju, D[39] | 2020 | Hefei, China | Prospective randomized, controlled study | INT=65.24±4.57; CON=65.83±4.08 | INT=9 (31.0); CON=14 (38.9) | Randomized=93; Completed=65; INT=29; CON=36 | Qigong | RT | 6 | Three times a week | PSQI |
| Lingling, Z[40] | 2020 | Shanghai, China | Prospective randomized, controlled study | AST=70.39±6.344; AT=69.75±6.195; CT=69.45±6.404 | AST=16 (28.1); AT=28 (46.7); CT=21 (37.5） | Randomized=173; Completed=173; AST=57; AT=60; CT=56 | CT | AST; AT | 1 | AST: once a day AT: three times a day | TE rate; PSQI |
| FengBin, Y et al. [41] | 2020 | Shanghai, China | Prospective randomized, controlled study | Age range=50-75 | None | Randomized=120; Completed=120; INT=60; CON=60 | AM | HE | 3 | Once a day | TE rate; PSQI |
| Yingqi, L[42] | 2020 | Heilongjiang, China | Prospective randomized, controlled study | INT=68.79±3.20; CON=68.26±3.50 | INT=15(46.9); C=17(50.0) | Randomized=72; Completed=66; INT=32; CON=34 | AAM | ACP | 1 | Six times a week | PSQI |
| Xin, W et al. [43] | 2020 | Heilongjiang, China | Prospective randomized, controlled study | INT=64.32±6.41; CON=63.42±7.01 | INT=13 (37.1); CON=10 (28.6) | Randomized=70; Completed=70; INT=35; CON=35 | CT | RT | 2 | Once a day | TE rate; PSQI |
| Bolun, S et al. [44] | 2020 | Changchun, China | Prospective randomized, controlled study | INT=63.47±1.64; CON=64.82±1.79 | None | Randomized=100; Completed=98; INT=49; CON=49 | MAM | HE | 0.5 | Three times a day | TE rate; PSQI |
| Zonghui, S et al. [45] | 2020 | Gansu, China | Prospective randomized, controlled study | INT=64.10±3.77; CON=65.23±4.85 | INT=14 (35.0); CON=12 (30.0) | Randomized=80; Completed=80; INT=40; CON=40 | CT | RT | 1 | Once a day | TE rate; PSQI |
| Hongshi, Z et al. [46] | 2019 | Jilin, China | Prospective randomized, controlled study | INT=57.63±3.90; CON=57.20±4.99 | INT=8 (26.7); CON=9 (30.0) | Randomized=60; Completed=60; INT=30; CON=30 | MT | RT | 1 | Once a day | TE rate; PSQI |
| Qiushuo, H et al. [47] | 2019 | Beijing, China | Prospective randomized, controlled study | Age range=35-70 | INT=9 (36.0); CON=11 (44.0) | Randomized=50; Completed=50; INT=25; CON=26 | CT | RT | 1 | Three times a week | TE rate; PSQI |
| Xueping, Y et al. [48] | 2019 | Heilongjiang, China | Prospective randomized, controlled study | INT=71.3±5.7; CON=72.3±4.8 | INT=15 (53.6); CON=12 (42.9) | Randomized=60; Completed=56; INT=28; CON=28 | ACP | RT | 1 | Once a day | TE rate; PSQI |
| Jiejun, H et al. [49] | 2019 | Shaanxi, China | Prospective randomized, controlled study | INT=65.50±5.30; CON=66.43±6.29 | INT=24 (52.2); CON=21 (47.7) | Randomized=90; Completed=90; INT=46; CON=44 | ACP | RT | 1 | Once a day | PSQI |
| Jinzhuo, F et al. [50] | 2019 | Hainan, China | Prospective randomized, controlled study | INT=68.45±5.30; CON=68.52±5.38 | INT=26 (52.0); CON=24 (49.0) | Randomized=99; Completed=99; INT=50; CON=49 | ACP | RT | 1 | Once a day | TE rate |
| Yongli, L et al. [51] | 2019 | Changchun, China | Prospective randomized, controlled study | INT=66.82±3.60; CON=67.31±3.03 | INT=18 (46.2); CON=20 (51.3) | Randomized=78; Completed=78; INT=39; CON=39 | CT | HMFB | 1 | Once a day | TE rate; PSQI |
| Yanwen, L[52] | 2019 | Guangdong, China | Prospective randomized, controlled study | INT=57.24±8.22; CON=56.90±7.92 | INT=18 (60.0); CON=17 (56.7) | Randomized=60; Completed=60; INT=30; CON=30 | AAM | ACP | 1 | Once every two days | TE rate; PSQI |
| Zhichao, Y et al. [53] | 2019 | Xiamen, China | Prospective randomized, controlled study | INT=60.4±8.35; CON=59.8±8.13 | INT=8 (40); CON=6 (30) | Randomized=40; Completed=40; INT=20; CON=20 | APAM | RT | 0.67 | Once a day | TE rate; PSQI |
| Yiming, C et al. [54] | 2019 | Foshan, China | Prospective randomized, controlled study | INT=74.98±12.32; CON=74.21±11.98 | INT=33 (57); CON=32 (56) | Randomized=114; Completed=114; INT=57; CON=57 | HMFB | RT | 1 | Once a day | TE rate; PSQI |
| Changmin, P et al. [55] | 2019 | Guangdong, China | Prospective randomized, controlled study | INT=51.07±7.23; CON=50.31±7.14 | INT=50(86); CON=50(98) | Randomized=50; Completed=50; INT=25; CON=25 | ACP | RT | 1 | Once a day | TE rate; PSQI |
| Juan, C et al. [56] | 2019 | xingtai, China | Prospective randomized, controlled study | INT=63±4; CON=62±3 | INT=30(58); CON=23(46) | Randomized=100; Completed=100; INT=51; CON=49 | ACP | RT | 0.67 | Once a day | TE rate |
| Donglie, W et al. [57] | 2019 | Beijing, China | Prospective randomized, controlled study | INT=66.76±3.58; CON=66.24±4.30 | INT=12(37.5); C=14(48.3) | Randomized=74; Completed=61; INT=32; CON=29 | Qigong | HE | 2 | Once a day | TE rate; PSQI |
| Yingwei, W et al. [58] | 2019 | Daqing, China | Prospective randomized, controlled study | INT=71.5±8.59; CON=72.31±8.63 | INT=21(46); CON=19(43) | Randomized=92; Completed=89; INT=45; CON=44 | Qigong | HE | 3 | Once a day | PSQI |
| Xuyu,Y et al. [59] | 2019 | Zhejiang, China | Prospective randomized, controlled study | INT=69.5±2; CON=70.5±3 | INT=34(44.2); CON=12(40.0) | Randomized=107; Completed=107; INT=77; CON=30 | ACP | RT | 1 | Once a day | TE rate; PSQI |
| Zhenzhen, J et al. [60] | 2018 | China | Prospective randomized, controlled study | ALL=71.6±8.1 | INT=60(58); C=60(59) | Randomized=120; Completed=120; I=60; C=60 | CT | HE | 1 | Once a day | TE rate; PSQI |
| Liaoqin, L et al. [61] | 2018 | Hunan, China | Prospective randomized, controlled study | INT=73.19±2.7; CON=73.16±2.71 | INT=17(45); C=17(45) | Randomized=74; Completed=74; INT=37; CON=37 | ACP | RT | 1 | Once a day | TE rate; PSQI |
| Fengzhi, G et al. [62] | 2018 | Beijing, China | Prospective randomized, controlled study | INT=61.60±6.007; CON=62.60±5.503 | INT=11(22); CON=6(12) | Randomized=100; Completed=100; INT=50; CON=50 | Qigong | RT | 2 | Once a day | TE rate; PSQI |
| Hong, Z et al. [63] | 2018 | Jiouquan, China | Prospective randomized, controlled study | ALL=69.7±1.2 | INT=30(53); CON=30(59) | Randomized=60; Completed=60; INT=30; CON=30 | AAM | HE | 0.47 | Once a day | TE rate |
| Wanhua, P et al. [64] | 2018 | Sandong, China | Prospective randomized, controlled study | Age range=60-80 | INT=20(51); CON=18(46) | Randomized=78; Completed=78; INT=39; C=39 | AT | RT | 1 | Once a day | TE rate |
| Fang, Y et al. [65] | 2018 | Beijing, China | Prospective randomized, controlled study | INT=66.7±5.75; CON=65.5±5.12 | INT=13(43); CON=14(46) | Randomized=120; Completed=120; INT=30; CON=30 | ACP | RT | 1 | Once a day | TE rate; PSQI |
| Jiangmei, L et al. [66] | 2018 | Kunming, China | Prospective randomized, controlled study | None | None | None | AAM | ACP | 2 | AAM: once a week; ACP: three times a week | TE rate; PSQI |
| Xiaohong, M et al. [67] | 2018 | Jiangsu, China | Prospective randomized, controlled study | INT=73.1±5.77; CON=75.23±4.85 | INT=23(76); CON=20(66) | Randomized=60; Completed=60; INT=30; CON=30 | CT | RT | 1 | Once a day | TE rate; PSQI |
| Yuting, T et al. [68] | 2017 | Hunan, China | Prospective randomized, controlled study | None | INT=42(53); CON=41(57) | Randomized=83; Completed=83; INT=42; CON=41 | CMIT | RT | 1 | Once a day | TE rate; PSQI |
| Yang, W et al. [69] | 2017 | Shenyang, China | Prospective randomized, controlled study | INT=51.47±8.32; CON=50.67±8.74 | INT=14(48); CON=16(55) | Randomized=58; Completed=58; INT=29; CON=29 | ACP | RT | 0.47 | Once a day | TE rate; PSQI |
| Hongjie, N et al. [70] | 2017 | Liaoning, China | Prospective randomized, controlled study | ALL=61.7±6.7 | INT=19(59); CON=18(56) | Randomized=64; Completed=64; INT=32; CON=32 | CT | RT | None | Once a day | TE rate |
| Wenxiong, X et al. [71] | 2017 | Zhangjiagang, China | Prospective randomized, controlled study | INT=69.01±2.15; CON=69.02±2.14 | INT=17(42); CON=18(45) | Randomized=80; Completed=80; INT=40; CON=40 | ACP | RT | 2 | Once every two days | TE rate; PSQI |
| Guifang, L et al. [72] | 2017 | Chengdu, China | Prospective randomized, controlled study | Age range=6-84 | INT=9(30); CON=8(26) | Randomized=92; Completed=90; INT=30; CON=30 | Moxibustion | RT | 1 | Once a day | TE rate; PSQI |
| Xiaomei, L et al. [73] | 2017 | Hangzhou, China | Prospective randomized, controlled study | Age range=60-75 | INT=14(49); CON=16(51) | Randomized=70; Completed=70; INT=35; CON=35 | AAM | RT | 1 | Once a day | TE rate; PSQI |
| Xuelian, Y et al. [74] | 2017 | Sichuan, China | Prospective randomized, controlled study | INT=71±5. 38; CON=71±5. 12 | INT=35(58); CON=33(55) | Randomized=120; Completed=120; INT=60; CON=60 | AM | HE | 1 | Once a day | TE rate; PSQI |
| Yuan, Z et al. [75] | 2016 | Bengbu, China | Prospective randomized, controlled study | Age range=60-86 | INT=12(45); CON=14(48) | Randomized=60; Completed=60; INT=30; C=30 | CHF | RT | 0.7 | Once a day | TE rate; PSQI |
| Wenjing, Z et al. [76] | 2016 | Shanghai, China | Prospective randomized, controlled study | INT=67.60±4.42; CON=68.35±5.10 | INT=15(30); CON=17(34.6) | Randomized=152; Completed=152; INT=50; CON=49 | CHF | RT | 4 | Once a day | TE rate; PSQI |
| Zongjun, M et al. [77] | 2016 | Kelamayi, China | Prospective randomized, controlled study | Age range=45-82 | INT=22(43.1); CON=18(35.3) | Randomized=102; Completed=102; INT=51; C=51 | AAM | ACP | 1 | Once a day | TE rate |
| Xuxing, L et al. [78] | 2016 | Fuzhou, China | Prospective randomized, patient-, analyst-blinded, controlled study | INT=65.63±5.44; CON=66.18±5.62 | INT=13(40.6); CON=8(26.7) | Randomized=105; Completed=95; INT=32; CON=30 | FEM | RT | 1 | Once a day | TE rate; PSQI |
| Feifei, W et al. [79] | 2016 | Hangzhou, China | Prospective randomized, controlled study | Age range=80-92 | ALL=52(65) | Randomized=80; Completed=80; INT=40; CON=40 | ACP | RT | 2 | None | PSQI |
| Heng, L et al. [80] | 2016 | Chengdu, China | Prospective randomized, controlled study | None | None | Randomized=60; Completed=56; AM=18; Aromatherapy=18; CON=20 | AM | Aromatherapy; RT | 2 | Twice a day | PSQI |
| Cunjv, Z et al. [81] | 2015 | Xuzhou, China | Prospective randomized, controlled study | Age=60-89 | None | Randomized=128; Completed=128; INT=64; CON=64 | HMFB | RT | 0.5 | Once a day |  |
| Jianping, W et al. [82] | 2015 | Shijiazhuang, China | Prospective randomized, controlled study | INT=73±6; CON=73±6 | INT=14(42.2); CON=12(40.0) | Randomized=98; Completed=98; INT=33; CON=30 | ACP | RT | 1 | Once a day | TE rate; PSQI |
| Yanfei, G et al. [83] | 2015 | Hangzhou, China | Prospective randomized, controlled study | INT=72.33±11.50; CON=71.67±12.00 | INT=31(47.7); CON=29(44.6) | Randomized=130; Completed=130; INT=65; CON=65 | CT | HMFB | 1 | Once a day | TE rate; PSQI |
| Xiandan, L et al. [84] | 2014 | Hangzhou, China | Prospective randomized, controlled study | Age range=65-80 | ALL=90(45) | Randomized=200; Completed=200; INT=100; CON=100 | CT | AT | 0.33 | Once a day | TE rate |
| Jun, H et al. [85] | 2013 | Hubei | Prospective randomized, controlled study | INT=59.25±15.65; CON=58.85±15.45 | INT=35(58.3); CON=39(65) | Randomized=120; Completed=120; INT=60; CON=60 | ACP | RT | 2 | Once a day in the first month, Once every other day in the second month | TE rate; PSQI |
| Yan, L et al. [86] | 2012 | Zhengzhou | Prospective randomized, controlled study | INT=66.48±6.59; CON=65.73±6.34 | INT=16(53.3); CON=19(63.3) | Randomized=60; Completed=60; INT=30; CON=30 | AT | RT | 1 | 4-5 times a day | TE rate; PSQI |
| Xinyan, L et al. [87] | 2010 | Beijing, China | Prospective randomized, controlled study | INT=59.50±8.11; CON=58.20±7.83 | INT=10(33.3); CON=12(40) | Randomized=60; Completed=60; INT=30; CON=30 | ACP | RT | 1 | Rest for 2 days,every 5 days of treatment | TE rate; PSQI |
| Zhaoyuan, S et al. [88] | 2010 | Tianjin, China | Prospective randomized, controlled study | INT=4.32±6.604; CON=4.48±6.687 | INT=22(55); CON=19(47.5) | Randomized=80; Completed=80; INT=40; CON=40 | ACP | RT | 1 | Once a day | TE rate; PSQI |
| Hong, C et al. [89] | 2009 | Shijiazhuang, China | Prospective randomized, controlled study | INT=66.15; CON=65.18 | INT=70(48.6); CON=64(48.5) | Randomized=276; Completed=276; INT=144; CON=132 | ACP | RT | 0.67 | None | TE rate |
| Shulan, Y et al. [90] | 2007 | Shenyang, China | Prospective randomized, controlled study | INT=68; CON=65 | INT=13(43.3); CON=10(35.7) | Randomized=58; Completed=58; INT=30; CON=28 | ACP | RT | 1 | Once a day | TE rate |
| Lianzhu, S et al. [91] | 2005 | Xingtai, China | Prospective randomized, controlled study | INT=65; CON=67 | INT=26(54.2); CON=44(47.9) | Randomized=96; Completed=96; INT=48; CON=48 | ACP | RT | 1.17 | Once a day | TE rate |
| Haili, S et al. [92] | 2002 | Beijing, China | Prospective randomized, controlled study | Age range=60-85 | ALL=96(64) | Randomized=150; Completed=150; INT=75; CON=75 | APAM | RT | 0.5 | Once a day | TE rate |
| Han, T et al. [93] | 2001 | Wuhu, China | Prospective randomized, controlled study | Age range=48-89 | ALL=85(44.97) | Randomized=189; Completed=189; AAM=63; ACP=63; AT=63 | AAM | ACP; AT | 1 | Once a day | TE rate |
| Xinling, T et al. [94] | 2010 | Weihai, China | Prospective randomized, controlled study | ALL=66.15 | INT=11(24.4); CON=9(20.0) | Randomized=90; Completed=90; INT=45; CON=45 | Moxibustion | RT | 1.5 | Once a day | TE rate |
| Chao, L et al[95] | 2010 | Shanghai, China | Prospective randomized, controlled study | INT=64.78±3.12; CON=64.69±3.10 | INT=24(48.0); CON=22(44.0) | Randomized=150; Completed=149; INT=50; CON=50 | Qigong | RT | 3 | Five times per week | TE rate |
| Wenjing, H et al.[96] | 2009 | Hubei, China | Prospective randomized, controlled study | ALL=75.15 | INT=25(83.3); CON=23(76.7) | Randomized=60; Completed=60; INT=30; CON=30 | HMFB | RT | 0.25 | Once a day | TE rate |
| Huabin, Z et al.[97] | 2008 | Chengdu, China | Multicenter randomized controlled trial | INT=70.32±4.236; CON=70.65±4.154 | INT=44(54.3); CON=39(49.4) | Randomized=241; Completed=241; INT=81; CON=79 | ACP | RT | 1 | Once a day | TE rate |
| Xinyan, L et al. [98] | 2008 | Beijing, China | Prospective randomized, controlled study | INT=59.50±8.11; CON=58.20±7.83 | INT=10(33.3); CON=12(40.0) | Randomized=60; Completed=60; INT=30; CON=30 | ACP | RT | 1 | Once a day | PSQI; TE rate |
| Hao, J et al. [99] | 2004 | Nanjing, China | Prospective randomized, controlled study | ALL=62 | INT=53(59.6); CON=25(62.5) | Randomized=129; Completed=129; INT=89; CON=40 | MT | RT | 1 | Twice a day | TE rate |
| Fengxian, L et al. [100] | 2021 | Guangxi, China | Prospective randomized, controlled study | INT=61.4±4.0; CON=62.0±3.8 | INT=30(69.8); CON=29(67.4) | Randomized=86; Completed=86; INT=43; CON=43 | AT | RT | 1 | Once a day | PSQI; TE rate |
| Lei, D et al. [101] | 2022 | Jiangsu, China | Prospective randomized, controlled study | None | INT=14(58.3); CON=11(45.8) | Randomized=48; Completed=48; INT=24; CON=24 | APAM | RT | 0.5 | Once a day | TE rate |
| Lin, G et al. [102] | 2021 | Anhui, China | Prospective randomized, controlled study | INT=73.41±6.48; CON=73.84±6.40 | INT=16(0.5); CON=15(46.8) | Randomized=64; Completed=64; INT=32; CON=32 | AAM | RT | 1 | Six times per week | TE rate; PSQI |
| Jiancheng, H et al. [103] | 2021 | Henan, China | Prospective randomized, controlled study | Age range=60-70 | None | Randomized=68; Completed=68; INT=34; CON=34 | Qigong | RT | 1.5 | Five times per week | TE rate; PSQI |
| Feifeng, W et al. [104] | 2022 | Nanjing, China | Prospective randomized, controlled study | INT=69.43±3.46; CON=70.35±2.84 | INT=19(63.3); CON=17(56.6) | Randomized=60; Completed=60; INT=30; CON=30 | CT | RT | None | None | TE rate; PSQI |
| Dongmei, Y et al. [105] | 2021 | Shandong, China | Prospective randomized, controlled study | INT=64.48±6.49; CON=63.49±6.47 | INT=22(44.8); CON=23(46.9) | Randomized=98; Completed=98; INT=49; CON=49 | AAM | ACP | 1 | Once every two days,30min each time | PSQI |
| Xinyan, L et al. [106] | 2021 | Ningxia, China | Prospective randomized, controlled study | INT=72.45±3.11; CON=71.10±2.48 | INT=21(46.6); CON=20(44.4) | Randomized=60; Completed=60; INT=30; CON=30 | ACP | RT | 1 | Once every two days | TE rate; PSQI |
| Lei, D et al. [107] | 2022 | Shandong, China | Prospective randomized, controlled study | INT=65.64±2.39; CON=65.42±2.45 | None | Randomized=94; Completed=90; INT=46; CON=44 | ACP | RT | 1 | Once every two days | TE rate |
| Ting, H et al. [108] | 2021 | Anhui, China | Prospective randomized, controlled study | INT=75.28±5.29; CON=75.64±5.37 | INT=30(55.56); CON=31(57.41) | Randomized=108; Completed=108; INT=54; CON=54 | HMFB | RT | None | None | TE rate |
| Xin, W et al. [109] | 2021 |  | Prospective randomized, controlled study | INT=65.42±5.38; CON=64.31±5.32 | None | Randomized=100; Completed=100; INT=50; CON=50 | AT | RT | None | None | TE rate |
| Giali, W et al. [110] | 2021 | Guangdong, China | Prospective randomized, controlled study | INT=67.01±5.12; CON=66.81±4.97 | INT=29(48); CON=30(51) | Randomized=119; Completed=119; INT=60; CON=59 | ACP | RT | 1 | Once a day | TE rate |
| Fan, B et al. [111] | 2020 | Shenzhen, China | Prospective randomized, controlled study | INT=70.3±5.7; CON=71.8±6.7 | ALL=34(24.5) | Randomized=139; Completed=129; INT=62; CON=57 | Qigong | RT | 7 | Five hours per week | PSQI |
| Yue, W et al. [112] | 2016 | Henan, China | Prospective randomized, controlled study | None | QAM=14(46.7); Qigong=13(43.3); CON=14(46.7) | Randomized=90; Completed=90; QAM=30; Qigong=30; CON=30 | QAM | Qigong; AM | 3 | Once a day | TE rate |
| Lai, F et al. [113] | 2017 | Taiwan, China | Randomized, controlled, single-blinded trial. | None | INT=12(38.7); CON=8(25.8) | Randomized=62; Completed=62; INT=31; CON=31 | AM | RT | 2 | Three times per week | PSQI |

Notes: INT: Intervention group; CON: Control group; ACP: Acupuncture; CT: Combination therapy; AAM: Acupuncture and moxibustion; AT: Auricular therapy; HE: Health education; HMFB: Herbal medicine foot bath; AM: Acupressure massage; APAM: Acupuncture plus acupressure massage; QAM: Qigong plus acupressure massage; MT: Manipulative therapy; CHF: Chinese herbal fumigation; MAM: Moxibustion plus acupressure massage; FEM: Five-element music; AST: Acupoint sticking therapy; CMIT: Chinese medicine iontophoresis treatment; TE rate: the total effective rate; PSQI: Pittsburgh Sleep Quality Index.

**Table 2** The total effective rate and Pittsburgh Sleep Quality Index rankings for different types of external treatments of traditional Chinese medicine

| Treatment | The total effective rate | | |  | Pittsburgh sleep quality index | | |
| --- | --- | --- | --- | --- | --- | --- | --- |
|  | SUCRA | Mean rank | P (%) |  | SUCRA | Mean rank | P (%) |
| RT | 7.4 | 16.7 | 0 |  | 4.3 | 18.2 | 0 |
| ACP | 45.9 | 10.2 | 0 |  | 36.2 | 12.5 | 0 |
| CT | 47.3 | 10 | 0 |  | 56.2 | 8.9 | 0 |
| Qigong | 78 | 4.7 | 1.3 |  | 64.7 | 7.4 | 0.2 |
| AAM | 76.6 | 5 | 2 |  | 87.4 | 3.3 | 17.5 |
| AT | 65 | 7 | 0.4 |  | 59.9 | 8.2 | 0.7 |
| HE | 12 | 16 | 0 |  | 16.4 | 16 | 0 |
| HMFB | 28 | 13.2 | 0 |  | 19.2 | 15.5 | 0 |
| AM | 75.4 | 5.2 | 1.7 |  | 90.9 | 2.6 | 32.9 |
| Moxibustion | 47 | 10 | 0.4 |  | 75.3 | 5.5 | 9.5 |
| APAM | 36.9 | 11.7 | 0 |  | 40.5 | 11.7 | 1.9 |
| QAM | 96.7 | 1.6 | 83.5 |  | — | — | — |
| MT | 51.5 | 9.2 | 0.6 |  | 59.1 | 8.4 | 7.3 |
| CHF | 15.3 | 15.4 | 0 |  | 18.6 | 15.7 | 0 |
| MAM | 62.9 | 7.3 | 4.8 |  | 54.4 | 9.2 | 6.1 |
| FEM | 61.7 | 7.5 | 3.6 |  | 51.7 | 9.7 | 3.8 |
| AST | 55.9 | 8.5 | 1 |  | 55 | 9.1 | 4.4 |
| CMIT | 36.3 | 11.8 | 0.7 |  | 57.9 | 8.6 | 9.4 |
| AST | 55.9 | 8.5 | 1 |  | — | — | — |
| CMIT | 36.3 | 0.7 | 11.8 |  | — | — | — |
| Aromatherapy | — | — | — |  | 55.5 | 9 | 3.7 |
| Scrapie | — | — | — |  | 46.8 | 10.6 | 2.6 |

Notes: Higher SUCRA and lower mean ranks indicate better-performing treatments. P (%) indicates the probability of it being the best treatment. ACP: Acupuncture; CT: Combination therapy; AAM: Acupuncture and moxibustion; AT: Auricular therapy; HE: Health education; HMFB: Herbal medicine foot bath; AM: Acupressure massage; APAM: Acupuncture plus acupressure massage; QAM: Qigong plus acupressure massage; MT: Manipulative therapy; CHF: Chinese herbal fumigation; MAM: Moxibustion plus acupressure massage; FEM: Five-element music; AST: Acupoint sticking therapy; CMIT: Chinese medicine iontophoresis treatment; SUCRA = surface under cumulative ranking curve.
